# A Comparative Analysis of Privacy-Preserving Large Language Models For Automated Echocardiography Report Analysis

**DOI:** 10.1101/2024.12.19.24319181

**Authors:** Elham Mahmoudi, Sanaz Vahdati, Chieh-Ju Chao, Bardia Khosravi, Ajay Misra, Francisco Lopez-Jimenez, Bradley J. Erickson

## Abstract

**Background:** Automated data extraction from echocardiography reports could facilitate large-scale registry creation and clinical surveillance of valvular heart diseases (VHD). We evaluated the performance of open-source Large Language Models (LLMs) guided by prompt instructions and chain of thought (CoT) for this task.

**Methods:** From consecutive transthoracic echocardiographies performed in our center, we utilized 200 random reports from 2019 for prompt optimization and 1000 from 2023 for evaluation. Five instruction-tuned LLMs (Qwen2.0-72B, Llama3.0-70B, Mixtral8-46.7B, Llama3.0-8B, and Phi3.0-3.8B) were guided by prompt instructions with and without CoT to classify prosthetic valve presence and VHD severity. Performance was evaluated using classification metrics against expert-labeled ground truth. Mean Squared Error (MSE) was also calculated for predicted severity’s deviation from actual severity.

**Results:** With CoT prompting, Llama3.0-70B and Qwen2.0 achieved the highest performance (accuracy: 99.1% and 98.9% for VHD severity; 100% and 99.9% for prosthetic valve; MSE: 0.02 and 0.05, respectively). Smaller models showed lower accuracy for VHD severity (54.1-85.9%) but maintained high accuracy for prosthetic valve detection (>96%). CoT reasoning yielded higher accuracy for larger models while increasing processing time from 2-25 to 67-154 seconds per report. Based of CoT reasonings, the wrong predictions were mainly due to model outputs being influenced by irrelevant information in the text or failure to follow the prompt instructions.

**Conclusions:** Our study demonstrates the near-perfect performance of open-source LLMs for automated echocardiography report interpretation with purpose of registry formation and disease surveillance. While larger models achieved exceptional accuracy through prompt optimization, practical implementation requires balancing performance with computational efficiency.

## Introduction

Content extraction from echocardiography reports is crucial for case retrieval and registry formation in cardiology. As enormous volumes of unstructured data are generated, the ability to rapidly extract and synthesize key clinical information has become essential for both individual patient care and population-level health research. Traditionally, this process has been manual, time-consuming, and prone to inconsistencies ^1^. Over the years, various natural language processing (NLP) techniques, have been proposed to automate and enhance this process ^2^.

The recent advent of Large Language Models (LLMs) has revolutionized the performance of AI models in text interpretation and generation, opening new avenues for medical report analysis ^3^. These models, trained on vast corpora of text data, learn to associate characters and words, enabling them to generate contextually appropriate responses. In medical applications, this translates to the ability to interpret complex clinical narratives and extract relevant information. However, the trade-off between model size, performance, and computational resources remains a critical consideration. Larger models, while more comprehensive, incur higher computational costs during both the training and the deployment ^4^.

To optimize LLM’s performance for specific tasks like echocardiography report interpretation, several strategies have been explored. Fine-tuning on domain-specific data has shown promise but is resource-intensive ^5^. On the other hand, many publicly released LLMs now offer instruction-tuned versions, specialized for tasks that require precise instruction following, which enables higher performance through prompt optimization, a simpler and more resource-efficient approach than domain-specific fine-tuning of a pretrained model ^6^. Patient data confidentiality is another concern in the analysis of medical report content ^7^. The rise of open-source LLMs has been particularly impactful in this regard. These models offer privacy-preserving solutions through on-premise deployment, addressing the critical concern of patient privacy ^8^.

There are reports on the use of open-source LLMs for translation or abstraction of medical reports, approaching or matching the performance of human experts ^9,10^. However, evidence on the performance of LLMs in free-text echocardiography report labeling is limited. In this study, we aim to develop an automated pipeline to label the echocardiography reports based on valvular heart diseases (VHD) severity and prosthetic valve presence. We utilized prompt engineering for a group of modern open-source LLMs with varying parameter sizes to assess trade-offs between computational complexity and abstraction accuracy. We also explore the effect of prompting strategy on model’s ability to extract structured information from the reports. Through the current survey, we aim to contribute to the growing body of knowledge on the application of language models in cardiovascular medicine, with a focus on improving the accessibility and utility of echocardiographic data for clinical and research purposes.

## Methods

### Data collection

In this Institutional Review Board (IRB) approved study (#24-006844), from 35,255 echocardiography reports generated during the year 2019, a sample of 200 reports were randomly selected for prompt optimization. From 33,058 echocardiography reports generated throughout the year 2023 a sample of 1000 reports were randomly selected for evaluation of our method’s generalizability to the current practice. Reports included two sections of “findings” and “impressions”. The impressions were a summarization of the findings’ content, hence excluded. The findings section of each report was collected as free-text content, which was often (yet not always), organized into five consistent categories: “Left Ventricle,” “Right Ventricle,” “Atria,” “Cardiac Valves,” and “Other Echocardiographic Findings.”

Reports were manually labeled by a cardiologist certified in echocardiography in 12 categories: prosthetic aortic, mitral, pulmonary and tricuspid valve, aortic regurgitation, aortic stenosis, mitral regurgitation, mitral stenosis, pulmonary regurgitation, pulmonary stenosis, tricuspid regurgitation, and tricuspid stenosis. Each prosthetic valve category was labeled as “yes” if a bioprosthetic or mechanical prosthetic valve was reported in the indexed valve’s position, or “no” if this condition was not satisfied. Each of the eight VHD categories was labeled as “absent”, “trivial”, “mild”, “mild to moderate”, “moderate”, “moderate to severe”, “severe”, or “not specified”. If the degree of VHD was not explicitly mentioned in the report it was labeled as “not specified”, while in the case of explicit mentioning of “no [VHD]“, the report was labeled as “absent” in that VHD category.

### Prompts structure

Prompts consisted of three main roles: system, user, and assistant. System role was defined as an expert cardiologist classifying the reports based on the grade or the presence of several conditions; User role contained a general instruction for the model to avoid either hallucination or the use of any pre-existing knowledge beyond the report’s content; It also included the report content followed by the assistant’s response to start the conversation. Two schemas were then imported presenting the 12 conditions (prosthetic valves and VHDs), predefined options (e.g., prosthetic, absent, mild, severe, etc.) and a condition-specific instruction, which could be followed by Chain of Thought (CoT) steps depending on a predefined parameter’s input to evaluate the pipeline in presence and absence of CoT reasoning. Prompt content structure is summarized in Figure 1 and a sample of a full prompt is provided in the supplementary material.

**Figure 1.**
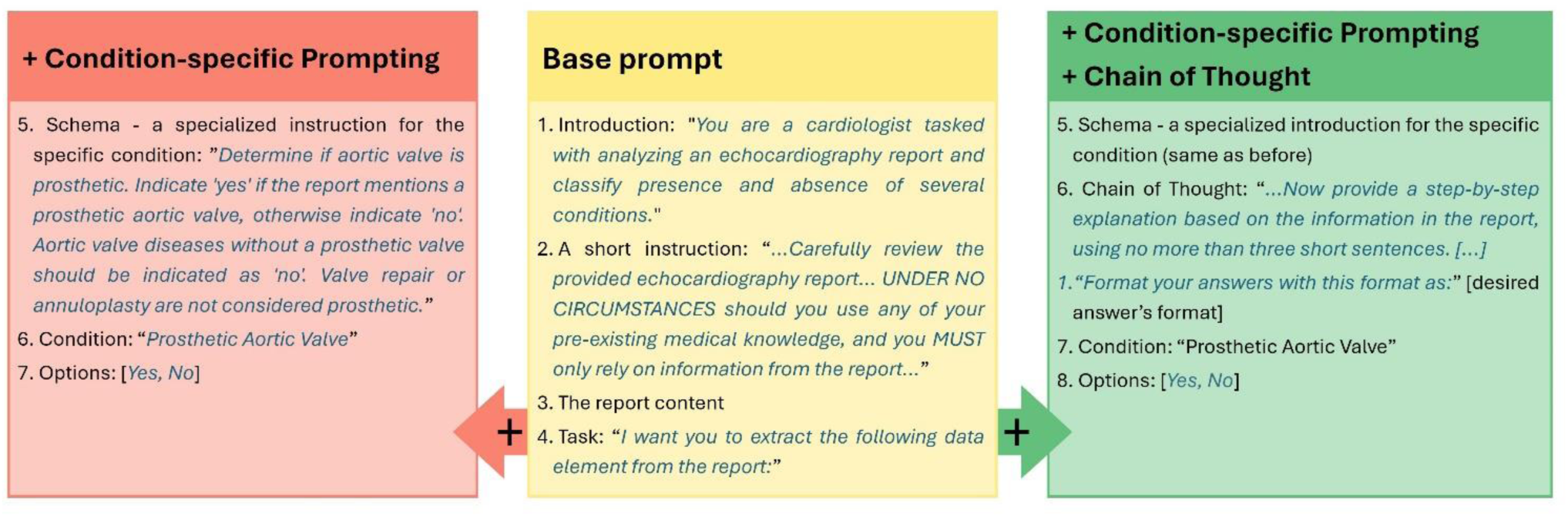
The prompt structure for prosthetic aortic valve classification: The base prompt (middle column) followed by condition-specific instructions - using two schema - alone (left column) or with chain of thought (right column).

Eventually, the models’ output was stored as a list of “condition”s (from Schema), “final answer” (selected from predefined options) and “explanation” (from CoT reasonings). The prompts were restarted after classifying each condition so that the model was blind to its responses to previous conditions.

### Model selection, and deployment

Five language models of the “Instruct” series were utilized for inference: Qwen2.0 (72.7B parameters) ^11^, Llama3.0 (70B parameters) ^12^, Mixtral8x7B (46.7B parameters) ^13^, Llama3.0 (8B parameters) ^12^, and Phi3.0-128K-mini (3.8B parameters). Parameter size, context window and token limit of the proposed models are summarized in Table 1. More details about the models and tokenization process are provided in supplementary material. All the codes were written in Python v3.9 and every model’s inference were run on a single NVIDIA A100-SXM4 80GB GPU. Prompt instructions and CoTs were utilized to elucidate the reasoning behind the interpretations of the language model. The final labels and the reasonings were recorded in tabular data frames format.

**Table 1.**
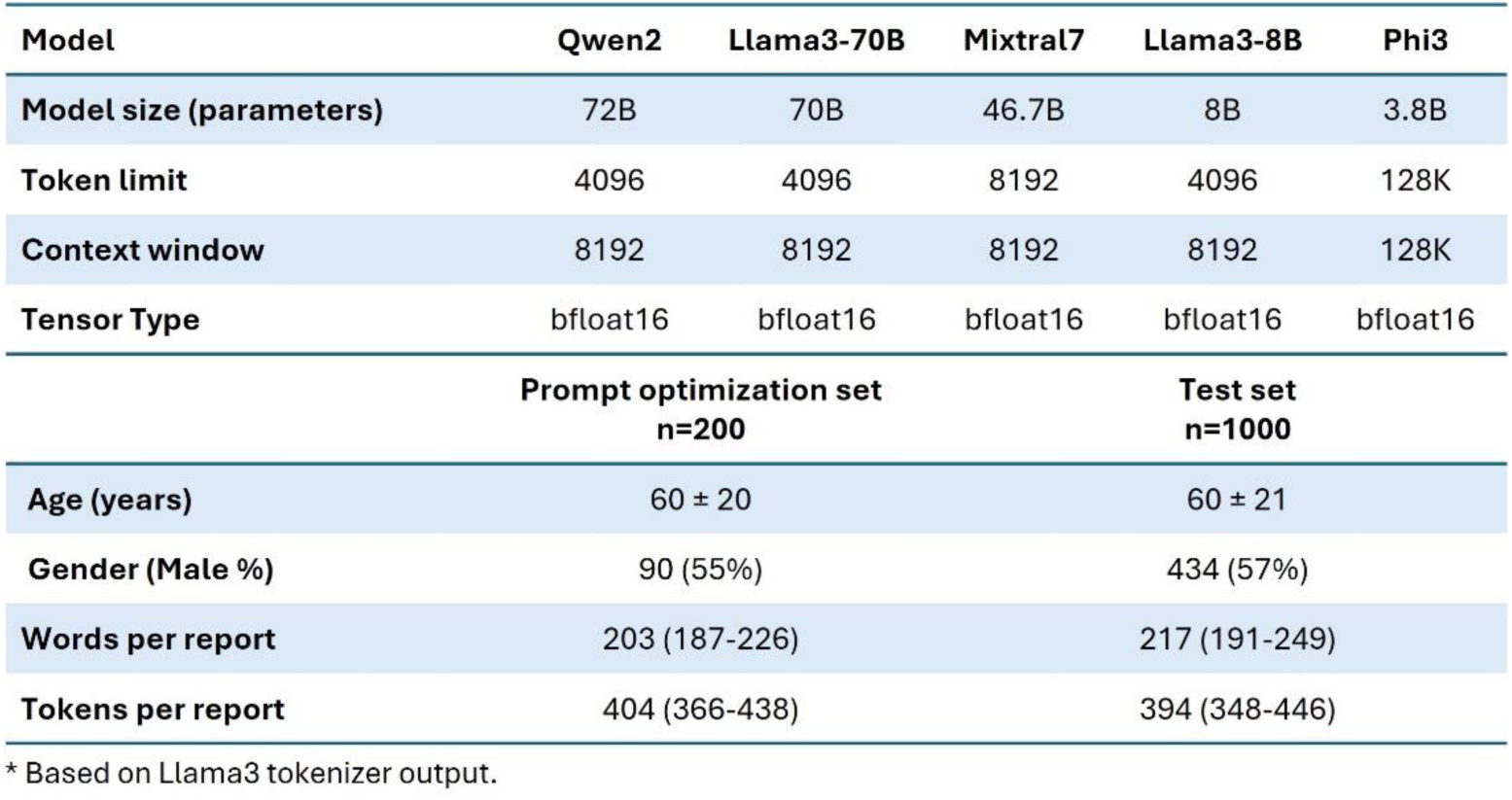
Proposed Language models and study population characteristics.

| Model | Qwen2 | Llama3-70B | Mixtral7 | Llama3-8B | Phi3 |
| --- | --- | --- | --- | --- | --- |
| Model size (parameters) | 72B | 70B | 46.7B | 8B | 3.8B |
| Token limit | 4096 | 4096 | 8192 | 4096 | 128K |
| Context window | 8192 | 8192 | 8192 | 8192 | 128K |
| Tensor Type | bfloat16 | bfloat16 | bfloat16 | bfloat16 | bfloat16 |
| Prompt optimization set<br>n=200 |  |  |  | Test set<br>n=1000 |  |
| Age (years) | $60 \pm 20$ | | | $60 \pm 21$ | |
| Gender (Male %) | 90 (55%) |  |  | 434 (57%) |  |
| Words per report | 203 (187-226) |  |  | 217 (191-249) |  |
| Tokens per report | 404 (366-438) |  |  | 394 (348-446) |  |

\* Based on Llama3 tokenizer output.

### Prompt optimization and model evaluation

Initially, each model’s inference was applied to the 200 reports from 2019. The incorrectly classified reports were investigated in detail by a certified cardiologist to detect the potential areas for prompt optimization. Based on the output CoT reasonings, the ‘hint’ parameter of the schemas was updated. This process was performed on each model independently based on its own incorrect classifications and reasonings. The inference was then reapplied to the reports and the iterations were continued until no improvement was seen in the model’s performance. The optimized pipeline was then tested on 1000 reports from 2023 to evaluate our pipeline’s performance on the echocardiography reports corresponding to the current practice.

No postprocessing was performed on the models’ output and the output labels were evaluated by sensitivity, specificity, positive and negative predictive values and accuracy; To explore the extent of the predictions’ error from the actual severities, the mean squared error (MSE) was calculated between the actual and predicted labels. To quantify the labels for MSE calculation, the VHD severities were recoded to a value from 1 to 7 for “absent” to “severe”; “not specified” was set to 0 since not mentioning the degree typically means that a significant VHD was not observed, so that classifying it as trivial or mild by LLM is less misleading that classifying it as sever.

## Results

### Data characteristics

Overall, 35,255 transthoracic echocardiography reports were extracted from 2019 and 33,058 from 2023, utilizing an institutional data retrieval platform. Patients’ characteristics are summarized in Table 1. The optimization (n=200) and evaluation sets (n=1000) included 55% and 57% male prevalence with a mean age of 60 ± 20 and 60 ± 21 years, respectively. Reports contained a median word count of 203 (187-226) and 217 (191-249), tokenized to 404 (366-438) and 394 (348-446) tokens per report, respectively.

The prevalence of prosthetic valves and VHD severities in study population are summarized in Supplementary Table 1. Aortic and mitral prosthetic valves were most common prosthetics, with prevalences of 10% and 3.5% in the optimization set, and 9.7% and 1.3% in the evaluation set, respectively. Pulmonary and tricuspid prosthetic valves were rare (≤1%). For VHD severity, valvular stenoses were predominantly “not specified” across datasets. The regurgitations were most commonly specified as “absent” (91/200 and 509/1000), followed by “trivial” (50/200 and 211/1000) for the aortic valve; “trivial” (105/200 and 569/1000), followed by “mild” (48/200 and 212/1000) for the mitral valve; “trivial” (110/200 and 639/1000), followed by “not specified” (41/200 and 218/1000) for pulmonary valve; and “trivial” (87/200 and 535/1000), followed by “mild” (68/200 and 258/1000) for tricuspid valve, in the optimization and evaluation sets, respectively (Figure 2).

**Figure 2.**
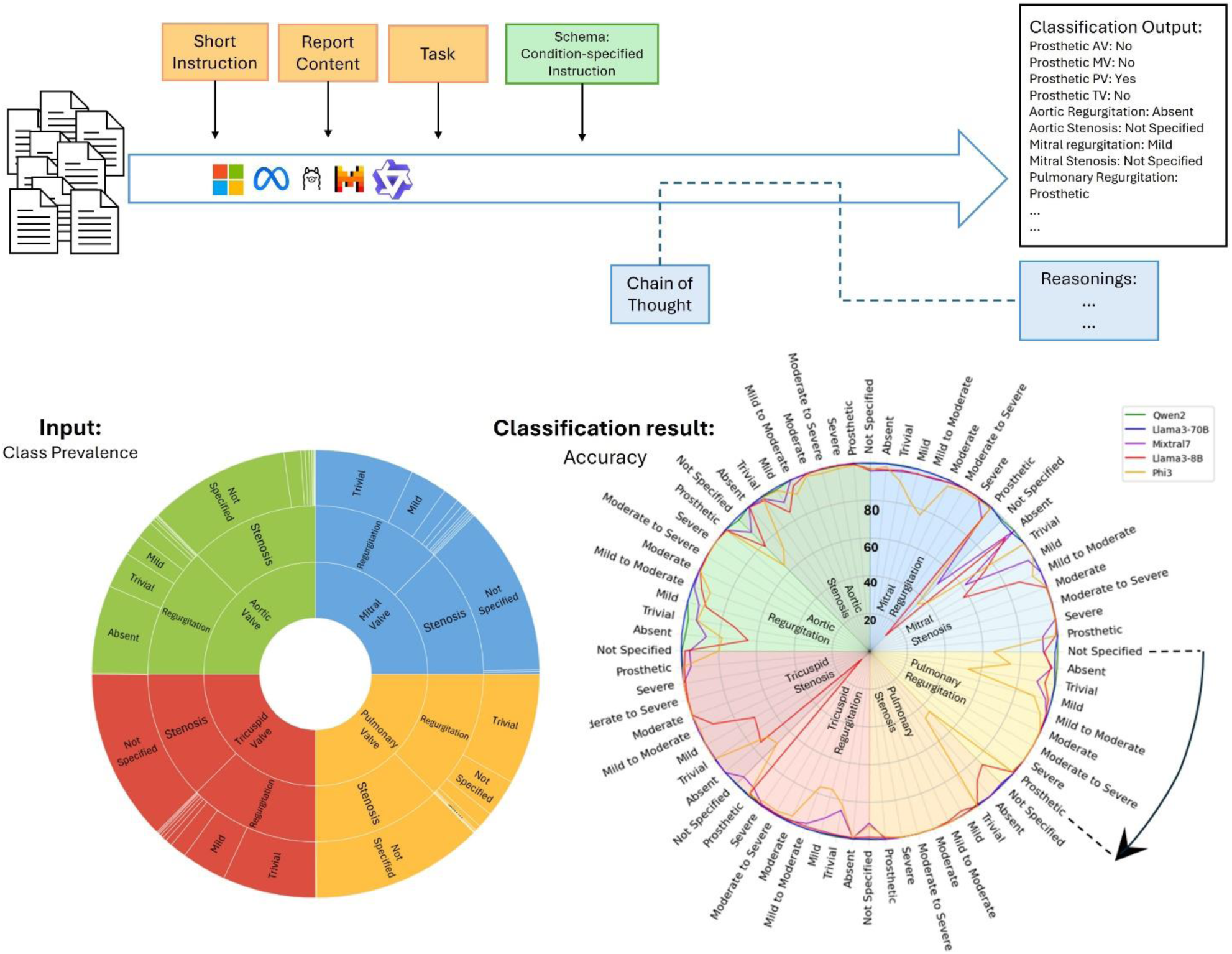
A schematic view of study workflow: Left - A sunburst plot of the distribution of severity classes for four valvular heart diseases. Right - A radar plot showing the accuracy of five proposed models in classification of nine categories of eight valvular heart disease severity (absent to severe and not specified) and prosthetic valve presence

In some VHD severities, like “absent mitral stenosis” or “moderate to severe pulmonary stenosis”, the positive cases were too few to conduct a reliable statistical evaluation; To overcome this limitation, the classification metrics were calculated only for categories labeled in more than 10 reports.

### Prompt Optimization

All five LLMs generated valid labels for 200 reports used in prompt optimization. At the end of the optimization, Qwen2.0, Llama3.0-70B, Mixtral8, Llama3.0-8B and Phi3.0 reached a mean accuracy of 98.6, 99.0, 85.9, 64 and 54.1 in severity labeling across all VHD categories and 100, 100, 95.5, 99.5 and 98.4 for prosthetic valves, respectively. Mean MSE reached to 0.14, 0.04, 0.64, 2.56 and 1.60, respectively. The evaluation metrics for all models per each category is provided in Supplementary Table 2.

### Evaluation

All five LLMs generated valid labels for 1000 reports used in evaluation. With CoT-guided prompting, Qwen2.0, Llama3.0-70B, Mixtral8, Llama3.0-8B and Phi3.0 yielded a mean accuracy of 98.9, 99.1, 85.9, 63.9 and 54.1 for VHD severity and 99.9, 100, 96.6, 99.0, 99.2 for prosthetic valves, respectively; Mean MSE was 0.05, 0.02, 0.57, 2.22 and 1.13, respectively. Without CoT-guided prompting, accuracies were generally lower, particularly for VHD severity classification. The exception was Phi3.0 which showed a lower accuracy and MSE in both tasks while guided by CoT prompting. Without CoT, Qwen2.0, Llama3.0-70B, Mixtral8, Llama3.0-8B and Phi3.0 yielded a mean accuracy of 94.0, 94.4, 81.1, 58.7 and 77.9 for VHD severity and 99.8, 99.9, 98.0, 96.3 and 99.8 for prosthetic valves, respectively; Mean MSE was 0.25, 0.09, 0.72, 2.69 and 0.73, respectively (Figure 3 and 4).

**Figure 3.**
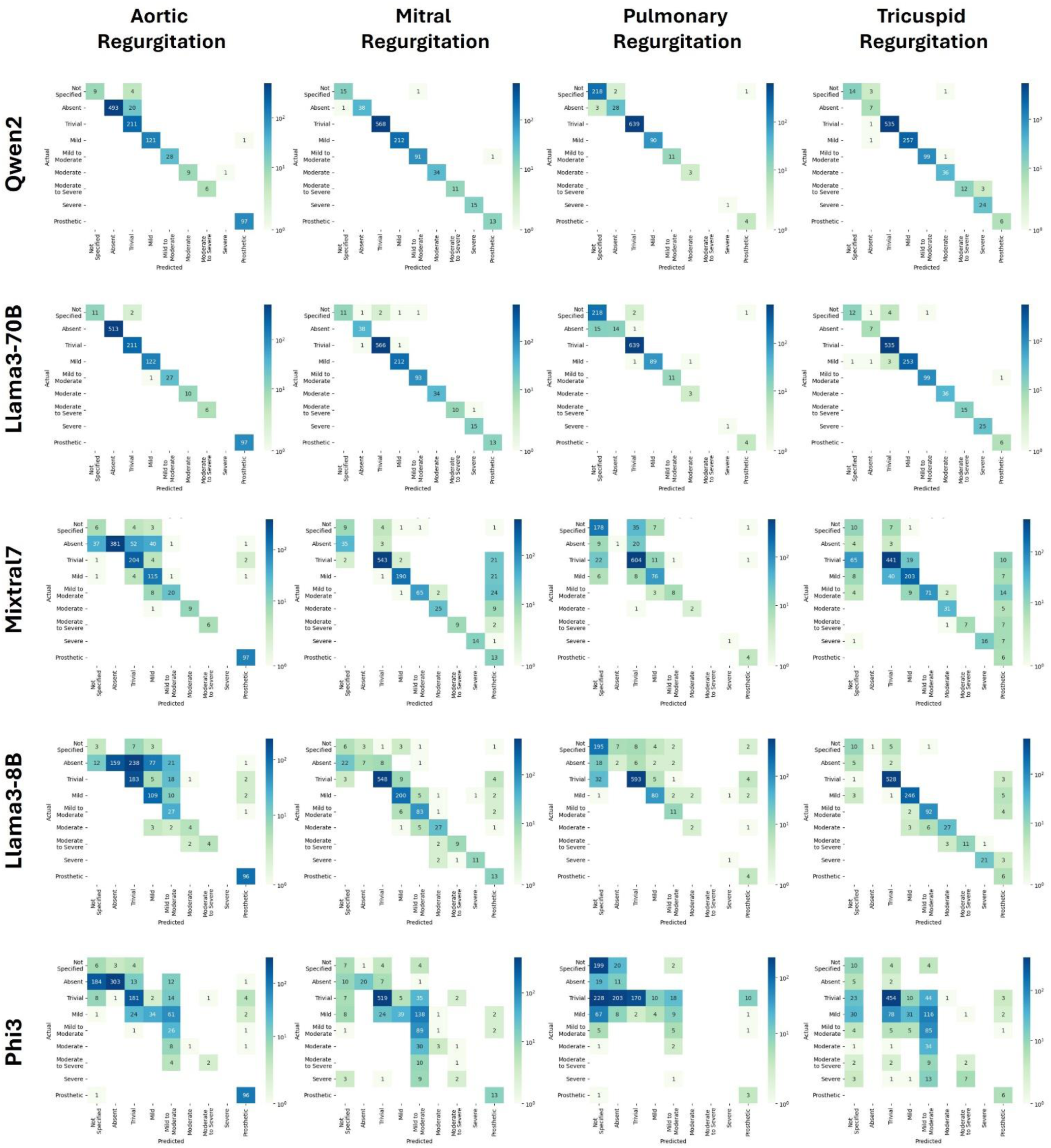
Heatmaps presenting five models’ predicted labels versus actual labels for four regurgitant valvular heart diseases.

**Figure 4.**
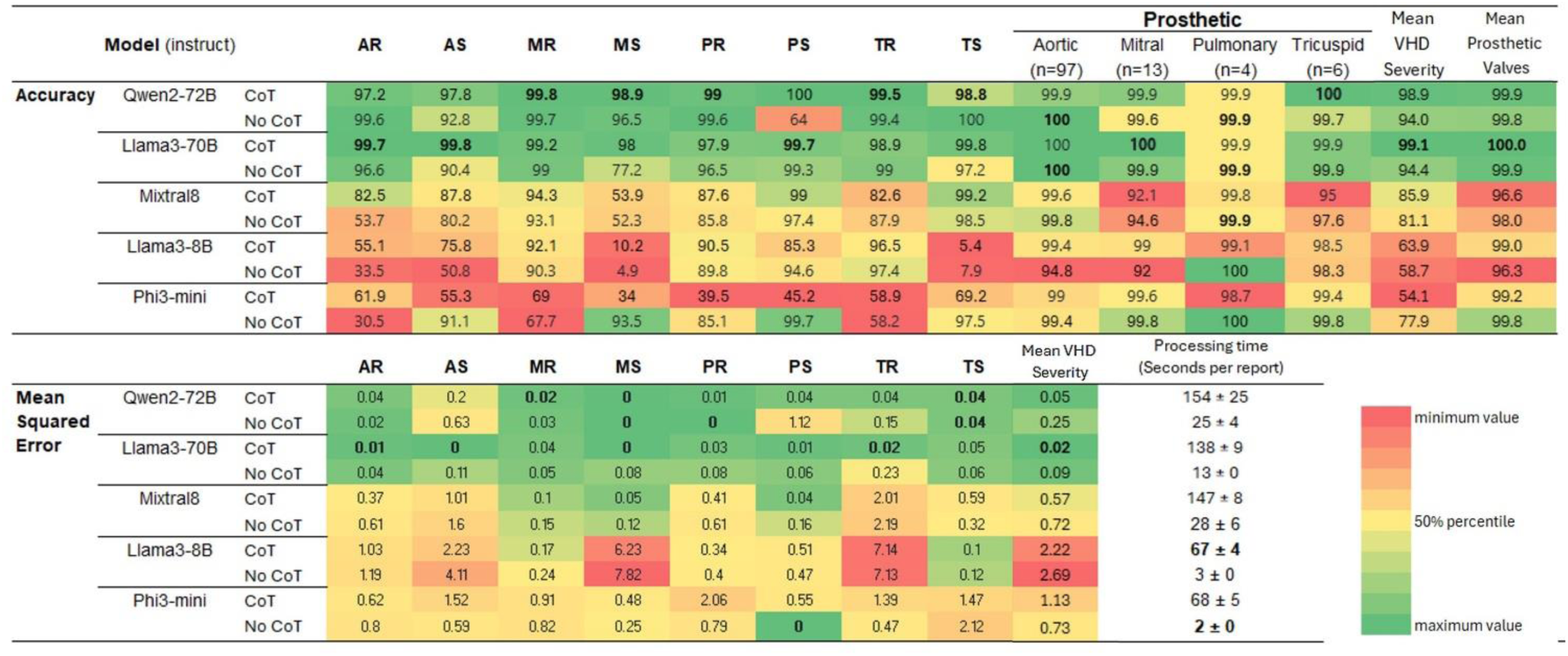
The processing time per report, the accuracy and mean squared error of each language model, with and without chain of thought, in classification of eight valvular heart disease categories, four prosthetic valve locations and across all diseases

According to the CoT reasonings of the wrong predictions, models’ wrong answers could be categorized in four general types which are exemplified in **Figure 4**: 1. Failure to detect the relevant data, which was rare by larger models including Qwen2.0, Llama3.0-70B; 2. Being influenced by irrelevant information, which was of the most common challenge for the larger models including Qwen2.0 and Llama3.0-70B; 3. Not following the prompt instructions, which was of the most common challenge for the larger models but happened in extreme prevalence for the smaller models like Phi3.0; 4. Generating nonfactual explanations (hallucinations), which almost exclusively happened for Phi3.0. It was not common for Llama3.0-8B and Mixtral8.

Notably, Mixtral8’s wrong severity predictions were close to the true labels and clinically reasonable in some cases, like considering thickened valves without specified stenosis as “trivial/mild stenosis”. On the other hand, Phi3.0 could not predict severity as “mild to moderate,” stating that this option is the safest or most fitting response, likely due to its position in the middle of the severity range. A more detailed analysis of each model’s potential sources of incorrect classifications is described in the supplementary material Section 2, with sample reports and reasonings provided.

**Table 2.** Six cases of ambiguous report content that could potentially interfere with the prompt instructions. Each model’s prediction is provided at the right columns with wrong labels written in red. AR: Aortic Regurgitation, MR: Mitral Regurgitation, PS: Pulmonary Stenosis, TR: Tricuspid Regurgitation.

| VHD | Report | Explanation | Expert | Final Answer |
| --- | --- | --- | --- | --- |
| Failure to detect the relevant data | “...Trileaflet aortic valve. Normal aortic valve. No aortic valve regurgitation. Mitral valve bowing. **No mitral valve regurgitation.** Normal pulmonary valve. Trivial pulmonary valve regurgitation. Normal tricuspid valve. Trivial tricuspid valve regurgitation. ...” | 1. The report mentions 'Mitral valve bowing' but does not specify the degree of mitral regurgitation.<br>2. The report does not mention 'mitral regurgitation' or 'MR'.<br>3. Therefore, the degree of mitral regurgitation is not specified. | Absent MR | Not Specified MR |
| Being influenced by irrelevant information | “...Trileaflet aortic valve. Thickened aortic valve. No aortic valve regurgitation. Thickened mitral valve. Trivial mitral valve regurgitation. Normal pulmonary valve. **Normal pulmonary valve systolic velocities. Trivial pulmonary valve regurgitation.** Normal tricuspid valve. Trivial tricuspid valve regurgitation...” | 1. The report mentions "Trivial pulmonary valve regurgitation."<br>2. The term "trivial" is used directly in relation to pulmonary valve function.<br>3. No other degree of pulmonary stenosis is mentioned. | Not Specified PS | Trivial PS |
| Not following the prompt instructions | “...Normal pulmonary valve. Trivial pulmonary valve regurgitation. **Status post [...] tricuspid valve annuloplasty.** Tricuspid valve diastolic mean Doppler gradient 4 mmHg (heart rate 92 BPM). Mild-moderate tricuspid valve regurgitation...” | 1. The report mentions "Status post [...] tricuspid valve annuloplasty" which suggests a prosthetic tricuspid valve.<br>2. The term "annuloplasty" implies a surgical repair or replacement of the valve. | No Prosthetic Tricuspid Valve | Prosthetic Tricuspid Valve |
|  | “...Trileaflet aortic valve. Normal aortic valve. No aortic valve regurgitation. Normal mitral valve. Trivial mitral valve regurgitation. Normal pulmonary valve. Trivial pulmonary valve regurgitation. **Normal tricuspid valve...**” | 1. The report mentions "Normal tricuspid valve."<br>2. There is no mention of regurgitation associated with the tricuspid valve.<br>3. The absence of any description of regurgitation implies that it is not present. | Not Specified TR | Absent TR |
| Confabulating nonfactual content | “...Bicuspid aortic valve. Aortic valve systolic mean Doppler gradient 25 mmHg. Aortic valve area by Doppler 2.38 cm2. **Moderate-severe aortic valve regurgitation.** Eccentric aortic valve regurgitant jet. Quantification with PISA was attempted but not reported. Normal mitral valve. Trivial mitral valve regurgitation. ...” | 1. The report mentions 'moderate aortic valve regurgitation'. 2. The term 'moderate' implies a degree between 'mild' and 'severe'. 3. Since 'moderate' is not an option, the closest appropriate category is 'mild to moderate'. Final Answer: mild to moderate | Moderate To Severe AR | Mild to Moderate AR |

Seven reports (from the whole 1200 dataset) contained ambiguous content that could potentially interfere with the prompt instructions. These reports and every model’s response to them is provided in Table 3. Notably, Qwen2.0 demonstrated superior performance in handling these ambiguities by consistently categorizing them as ‘not specified,’ which aligns with the approach used during ground truth labeling.

**Table 3.** Six cases of ambiguous report content that could potentially interfere with the prompt instructions. Each model’s prediction is provided at the right columns with wrong labels written in red. AR: Aortic Regurgitation, MR: Mitral Regurgitation, PR: Pulmonary Regurgitation.

| Ambiguous Reports |  | Valvular Heart Disease | Ground Truth | Qwen2 | Llam3-70B | Mixtral7 | Llam3-8B | Phi3 |
| --- | --- | --- | --- | --- | --- | --- | --- | --- |
| Severity degree was not mentioned. | "... Bicuspid aortic valve with fused right and non coronary cusps. Thickened aortic valve. **Aortic valve regurgitation**. Diastolic flow reversals in the descending thoracic aorta. Diastolic flow reversals in the abdominal aorta. Thickened mitral valve. Trivial mitral valve regurgitation..." | AR | Not Specified | Trivial | Trivial | Trivial | Trivial | Trivial |
|  | "... Sclerotic mitral valve. Calcified mitral annulus. Mild mitral valve regurgitation. Normal pulmonary valve. Normal pulmonary valve Doppler flow pattern. **Pulmonary valve regurgitation**. Normal tricuspid valve. mild to moderate tricuspid valve regurgitation..." | PR | Not Specified | Not Specified | Not Specified | Trivial | Mild | Mild to Moderate |
|  | "...Trileaflet aortic valve. Normal aortic valve. **Aortic valve regurgitation**. Normal mitral valve. Trivial mitral valve regurgitation. Normal pulmonary valve. Trivial pulmonary valve regurgitation. Normal tricuspid valve. Trivial tricuspid valve regurgitation..." | AR | Not Specified | Not Specified | Trivial | Trivial | Trivial | Not Specified |
|  | "...Trileaflet aortic valve. Thickened aortic valve. No aortic valve regurgitation. Thickened mitral valve. **Eccentric mitral valve regurgitation** directed posteriorly (main regurgitant jet). Mitral regurgitation ERO (PISA) 0.15 cm2. Mitral regurgitant volume (PISA) 33 mL. Normal pulmonary valve. Normal pulmonary valve systolic velocities. Trivial pulmonary valve regurgitation. Normal tricuspid valve. Trivial tricuspid valve regurgitation..." | MR | Not Specified | Mild to Moderate | Absent | Mild to Moderate | Mild | Trivial |
| An unusual term was used to describe the degree of severity. | "...Trileaflet aortic valve. Sclerotic aortic valve. Aortic valve systolic mean Doppler gradient 5 mmHg. Mild aortic valve regurgitation. Thickened mitral valve. **Indeterminate degree of mitral valve regurgitation**. Mitral valve diastolic mean Doppler gradient 2 mmHg (heart rate 52 BPM). Normal pulmonary valve. Normal pulmonary valve systolic velocities. No pulmonary valve regurgitation. Normal tricuspid valve. Mild tricuspid valve regurgitation..." | MR | Not Specified | Not Specified | Mild | Not Specified | Prosthetic | Not Specified |
|  | "...Trileaflet aortic valve. Thickened aortic valve. **Aortic valve regurgitation (trace amount)**. Normal mitral valve. Trivial mitral valve regurgitation. Normal pulmonary valve. Normal pulmonary valve systolic velocities. Trivial pulmonary valve regurgitation. Normal tricuspid valve. Trivial tricuspid valve regurgitation..." | AR | Trivial | Trivial | Trivial | Trivial | Trivial | Trivial |
| Report did not include typical sections. | "Status post [...] procedure (e/w, ...). [...] procedure with valve sparing [...] Hemashield ascending and aortic root repair and coronary artery reimplantation (e/w, ...). ..." | AR | Prosthetic | Prosthetic | Prosthetic | Prosthetic | Prosthetic | Not Specified |
|  |  | MR | Not Specified | Not Specified | Not Specified | Not Specified | Not Specified | Not Specified |

## Discussion

Our study presents an efficient strategy for data extraction from echocardiography reports using open-source LLMs. We evaluated five models - Qwen2.0(72B), Llama3.0 (70B), Mixtral8, Llama3.0 (8B), and Phi3.0 - using prompt instructions and CoT reasoning. The larger models, Llama3.0 (70B) and Qwen2.0 (72B), achieved exceptional performance, with specificities of 99-100% and sensitivities of 98-100% across all VHD categories, without requiring domain-specific fine-tuning - a significant advantage for resource efficiency and generalizability. While model size correlated with performance, it was not the sole determinant, as evidenced by similar performance between Qwen2.0 (72B) and Llama3.0 (70B) despite Qwen2.0’s reported superiority in various benchmarks ^11^.

The high accuracy of these models enables the creation of comprehensive VHD registries with numerous clinical applications: automated registry creation reduces manual effort ^1^ allowing clinicians to focus more on patient care; standardized severity classification enables consistent disease tracking ^14^ and the ability to rapidly process historical reports enhances research potential. This can facilitate the development of automated alerting systems that flag significant changes in VHD severity leading to timely clinical review and adjustment of management strategy.

Previous studies have explored LLMs for echocardiography report interpretation through various approaches. While some achieved promising results using fine-tuned models for report summarization ^5^ or longitudinal analysis ^14^, our study demonstrated superior performance in both intervention identification and disease severity classification through prompt optimization and CoT reasoning. Due to the complexity of free-text content comparison, previous studies mainly evaluated their free-text output by natural language generation metrics (e.g., BLEU, METEOR and BERT score), which quantify the phrase and word similarities rather than the concept or meanings or based on expert’s selection of the “winner” or “correct” answer, which subjects the evaluator to confirmation bias leading to overestimation of model’s performance. We employed predefined output labels enabling objective evaluation through classification metrics. (e.g., sensitivity/recall, specificity, predictive values and accuracy). This approach has been previously proposed in traditional NLP studies ^15,16^.

Our observation of consistently high accuracy (>97.9%) across all VHD categories with Llama3.0-70B demonstrates a marked improvement over traditional NLP techniques, which have shown wide ranges of accuracies in similar tasks: from 45% for stenotic VHDs to 98% for mitral regurgitation ^17^, positive and negative predictive values of 99% and 96% for aortic stenosis ^15^, and recalls of 91-95% for VHD severity ^16^. The consistency of our results across all valve types and severity grades represents a significant advancement over these earlier approaches. This aligns with findings from other medical domains, including radiology report analysis ^18–20^.

This advanced performance rises from two key factors: the open-source LLMs are trained on a comprehensive corpus of data, giving them a basic knowledge of medical conditions, enabling them to interpret the concepts rather than rule-based text identification or limited embedding of similar inputs (exemplified by accurate classification of “trace” regurgitation as “trivial”); and the ability of instruction-tuned LLMs to interpret and address prompt instructions, further enabling them to minimize misinterpretations and invalid output generation.

However, LLMs have inherent limitations that warrant careful usage and validation of their outputs. As evident in our experiments, hallucination was a major weakness. While lower temperatures and concrete prompting ^21^ can reduce hallucination, the effect of model size on hallucination vulnerability is complex. Larger models, due to their more comprehensive knowledge base, are prone to generate irrelevant answers while being questioned about concepts absent from the input prompt (e.g., “who is the president of the United States?” or “When did the first aortic valve replacement perform?”). On the other hand, they are less likely to misinterpret information explicitly present in input prompts and this better interpretation of prompt content, including the instructions, makes them more steerable by prompt instructions ^22^, particularly after instruction-tuning.

This aligns with our findings where larger models achieved more accurate label extraction, while intermediate-sized models like Mixtral 8.0 and Llama 3.0 (8B), despite superior severity classification, underperformed in prosthetic valve identification compared to our smallest model, Phi3.0-mini; they persisted in misclassifying non-replacement valvular procedures as prosthetic valve replacement, even when explicitly instructed otherwise. A possible explanation is that intermediate-sized models occupy a problematic middle ground: they possess sufficient knowledge to recognize various valvular procedures but lack the robust instruction-following capabilities of larger models to prevent them from generating incorrect labeling of different procedures in category of prosthetic valve replacement.

The second challenge in application of LLMs involves the trade-off between accuracy and computational efficiency. While CoT reasoning substantially improved performance, it increased processing time from 2-25 seconds to 67-154 seconds per report. This trade-off becomes particularly relevant for real-time clinical applications. Future implementations may need to balance these factors through selective application of CoT reasoning or parallel processing strategies, though optimal solutions will likely be task-specific and require further validation.

Eventually, the very low prevalence of stenotic VHDs in the studied population, was a limitation of our randomly selected sample. Case retrieval from a large registry of labeled echocardiography reports is needed for further validation on rare VHD categories.

## Conclusion

In conclusion, our study demonstrates the significant potential of open-source LLMs in automating echocardiography report interpretation with purpose of registry formation, case retrieval and patient surveillance. The high accuracy achieved by models like Llama3.0 (70B) and Qwen2.0 (72B), coupled with the benefits of prompt instructions and CoT reasoning, represents a promising step towards more efficient and accurate cardiac imaging analysis. As these technologies continue to evolve, their integration into clinical workflows could substantially enhance decision-making processes and research capabilities in cardiology. However, it’s crucial to approach this integration thoughtfully. While LLMs show great promise, they should be viewed as supportive tools rather than replacements for clinical expertise. Ongoing validation, careful implementation, and continuous monitoring will be essential to harness the full potential of these models while ensuring patient safety and care quality.

## Supporting information

Supplementary methods and results

## Data Availability

All data produced in the present study are available upon reasonable request to the authors

## Supplementary materials

## 1. Methods

### Schema’s structure and content

To organize the content of the prompts, two schemas were developed. The first one contained four conditions of “Aortic Prosthetic Valve”, “Mitral Prosthetic Valve”, “Pulmonary Prosthetic Valve” and “Tricuspid Prosthetic Valve; Each condition was associated with two options of “yes” or “no”. The second one contained eight conditions for eight VHDs (e.g., aortic stenosis, aortic regurgitation, mitral stenosis, etc.), and 8 severity options (absent to severe and not specified) for each condition. Every condition in these two schemas also contained a “hint” prompt containing a condition-specific instruction to guide the model in its selection process. As an instance, a part of the hint for mitral stenosis was: ‘The report may use “MS” or “mitral stenosis” instead of “mitral valve stenosis”. “Normal mitral valve” without explicitly mentioning the degree of stenosis, does not mean “no MS”. ‘

### Sample of a full prompt

[**System**] You are a cardiologist tasked with analyzing an echocardiography report and classify presence and absence of several conditions.

[**User**] Carefully review the provided echocardiography reports (between three backticks). Ensure that each data element is accurately captured. Your attention to detail is crucial for maintaining the integrity of the medical records. You should not confabulate information, and if something is not mentioned, you should assume that it is ’not mentioned’ unless otherwise stated. UNDER NO CIRCUIMSTANCES you should use any of your pre-existing medical knowledge, and you MUST only rely on information from the report. We are interested at findings at the time of scan, not the previous ones, so only consider the impression and findings sections of the report.

’’’report

PROCEDURE: Transthoracic Echocardiogram

INDICATION: Evaluation of cardiac function.

FINDINGS: The left atrium is mildly dilated. The left ventricle has normal wall thickness and normal systolic function with an ejection fraction of 65%. There is no aortic regurgitation. The mitral valve shows mild to moderate regurgitation. The right ventricle has normal size and function. The tricuspid valve demonstrates trivial regurgitation. The pulmonary valve is normal.

IMPRESSION: 1. Normal left ventricular systolic function 2. No aortic regurgitation 3. Mild to moderate mitral regurgitation 4. Trivial tricuspid regurgitation 5. Normal pulmonary valve

’’’

Are you ready?

[**Assistant**] Yes! Send me the data elements that you want extracted from the report. When you ask me for explanation, I will only reason and when you ask me for the final answer, I will only provide the final answer. I will keep my responses short and to the point.

[**User**] I want you to extract the following data element from the report:

’Prosthetic Aortic Valve’

Here are your options and you can explicitly use one of these:

- ’yes’
- ’nò

Hint: Determine if aortic valve is prosthetic. Indicate ‘yes’ if the report mentions a prosthetic aortic valve, otherwise indicate ‘no’. Aortic valve diseases without a prosthetic valve should be indicated as ‘no’. Valve repair or annuloplasty are not considered prosthetic.

After you provide the data element, I will ask you to provide an explanation and then the final answer.

Now give your initial answer. Then provide a step-by-step explanation based on the information in the report, using no more than three short sentences. You can use fewer sentences if needed. Try to critically appraise your initial answer, which MIGHT be wrong. Then give me your final answer. Format your answers with this format as:

’’’answer

Initial Answer:

<YOUR initial answer>

Explanation:

1. <YOUR first step>
2. <YOUR second step> (if applicable)
3. <YOUR third step> (if applicable)

Final Answer:

<YOUR final answer>

’’’

**[the same for four prosthetic valves]**

[**User**] I want you to extract the following data element from the report:

’Aortic Regurgitation’

Here are your options and you can explicitly use one of these:

- ’absent’
- ’trivial’
- ’mild’
- ’mild to moderatè
- ’moderatè
- ’moderate to severè
- ’severè
- ’not specified’

Hint: The report may use ‘AR’ or ‘aortic regurgitation’ instead of ‘aortic valve regurgitation’. If the regurgitation degree is not explicitly mentioned in the report, indicate ‘not specified’ and do not indicate any other degree class. Regurgitation of other valves does not mean aortic valve necessarily has a regurgitation; AR can be ‘not specified’, ‘absent’ or any other class in the presence of other valves regurgitation. Indicate ’absent’ only if the report explicitly mentions the patient has ‘no’ aortic regurgitation. Do not indicate any other degree if the report explicitly mentions ‘no aortic regurgitation’. If the report mentions ‘trivial aortic regurgitation’, indicate ‘trivial’ and do not consider it ‘mild’.

After you provide the data element, I will ask you to provide an explanation and then the final answer.

Now give your initial answer. Then provide a step-by-step explanation based on the information in the report, using no more than three short sentences. You can use fewer sentences if needed. Try to critically appraise your initial answer, which MIGHT be wrong. Then give me your final answer. Format your answers with this format as:

’’’answer

Initial Answer:

<YOUR initial answer>

Explanation:

1. <YOUR first step>
2. <YOUR second step> (if applicable)
3. <YOUR third step> (if applicable)

Final Answer:

<YOUR final answer>

’’’

**[the same for eight valvular heart diseases]**

### Inference parameters

Sampling was deterministic, so the temperature, top-k and top-p selections were kept as models’ default. To optimize computational efficiency while maintaining model performance, we employed 4-bit quantization using the BitsAndBytes library and local caching was used for model loading and inference. The models and the corresponding tokenizers were loaded by the Hugging Face’s “Transformers” package. The chat pipeline and templates, output generation, and class selection were performed by Microsoft’s “Guidance” package.

## 2. Results

### Llama3-Instruct (70B)

Llama3-70B showed the best performance with an accuracy of 99.1% for VHD severities and 100% for prosthetic valve presence. It successfully classified all but a few cases in all categories. The false classifications were mainly for being influenced by irrelevant information; generating nonfactual explanations, absent from the prompt input, was very rare. There were also a few cases of classifying “mild to moderate” as “mild” or “moderate to severe” as “severe”. A case of tricuspid valve annuloplasty and a case of pulmonary valve homograft for aortic valve replacement were falsely classified as tricuspid and pulmonary valve prosthetic; otherwise, the prosthetic valves were classified correctly.

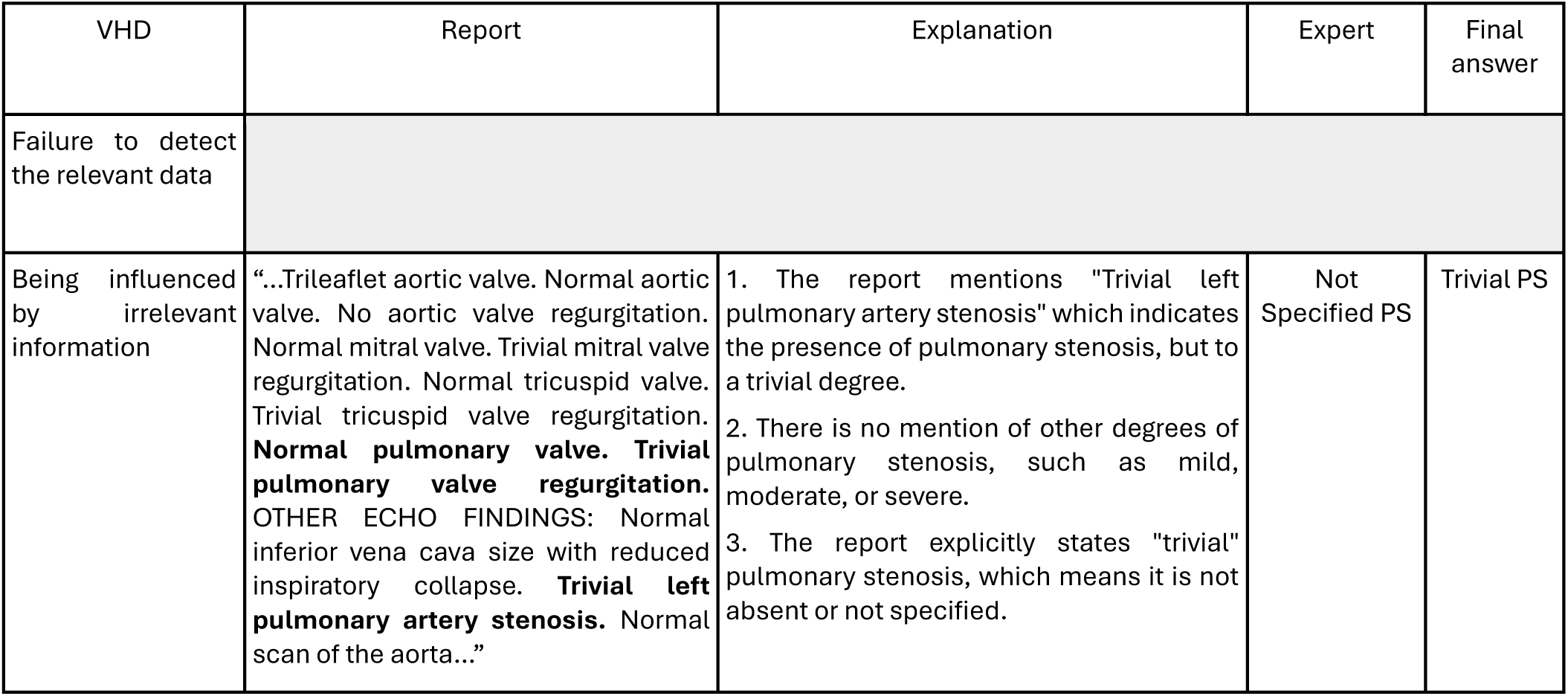
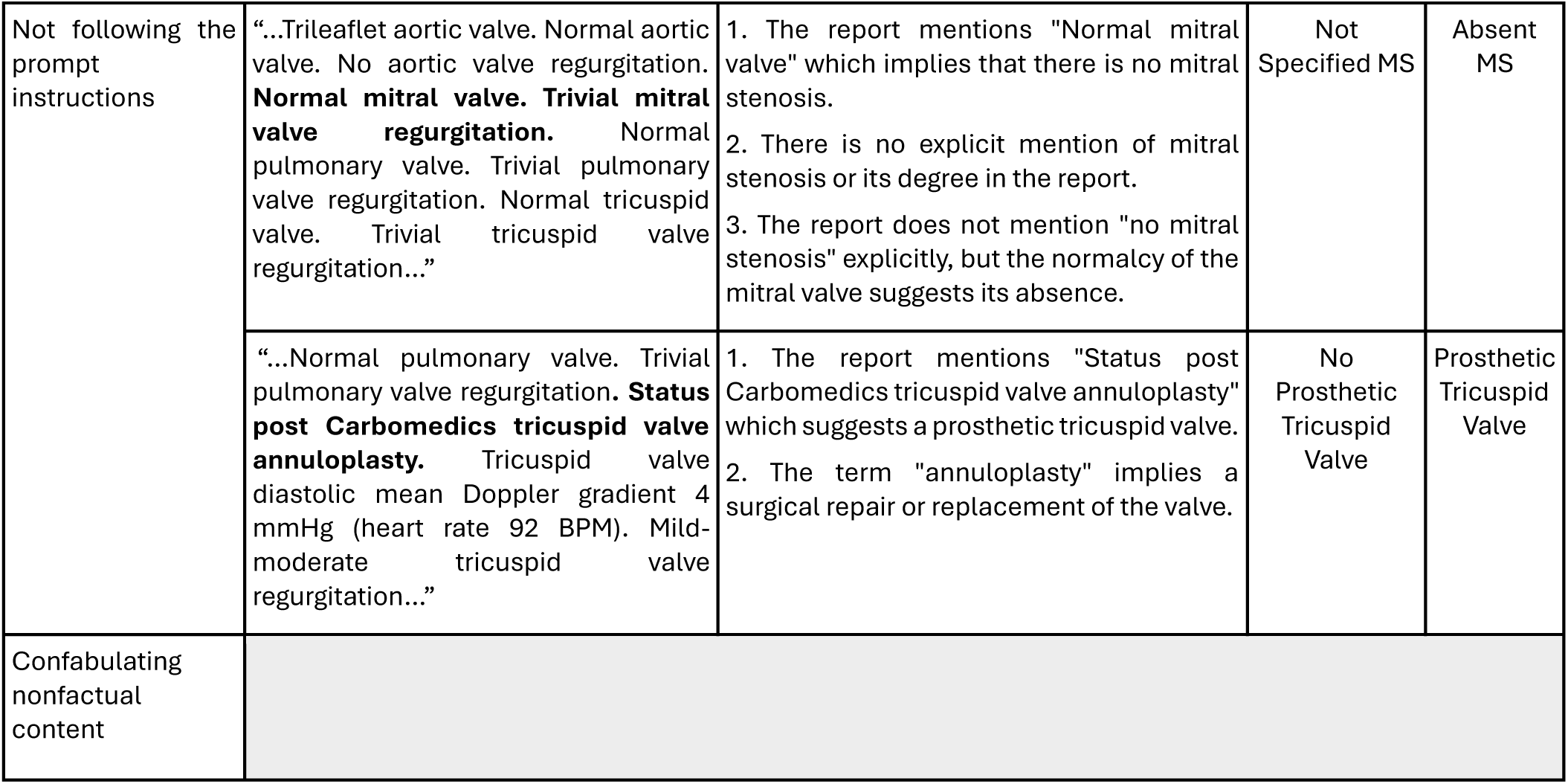

### Qwen2-Instruct (72.7B)

Qwen2 showed the second-best performance with an accuracy of 98.9% in the classification of VHD severities and 99.9% in the classification of prosthetic valve presence. Its most common mistake was to be influenced by irrelevant information in the report, rather than classifying the VHD degree based on the explicit degree mentioned. An example was classifying the sclerotic or thickened aortic valves as “mild AS”, while the degree of stenosis was not explicitly mentioned throughout the report and this concept was particularly clarified for the LLM in the prompts. This led to an accuracy of 97% in the classification of AR and AS. Qwen2 had the same problem of classifying moderate to severe as severe in a few cases and false classification of an aortic, a mitral, and a pulmonary repair as prosthetic valve.

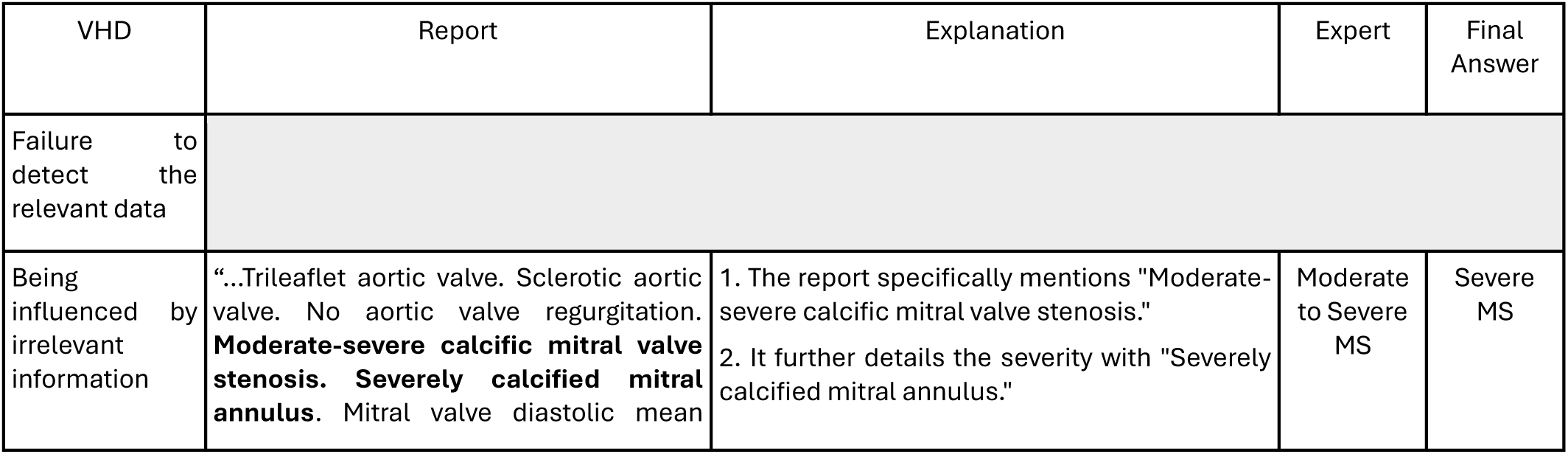
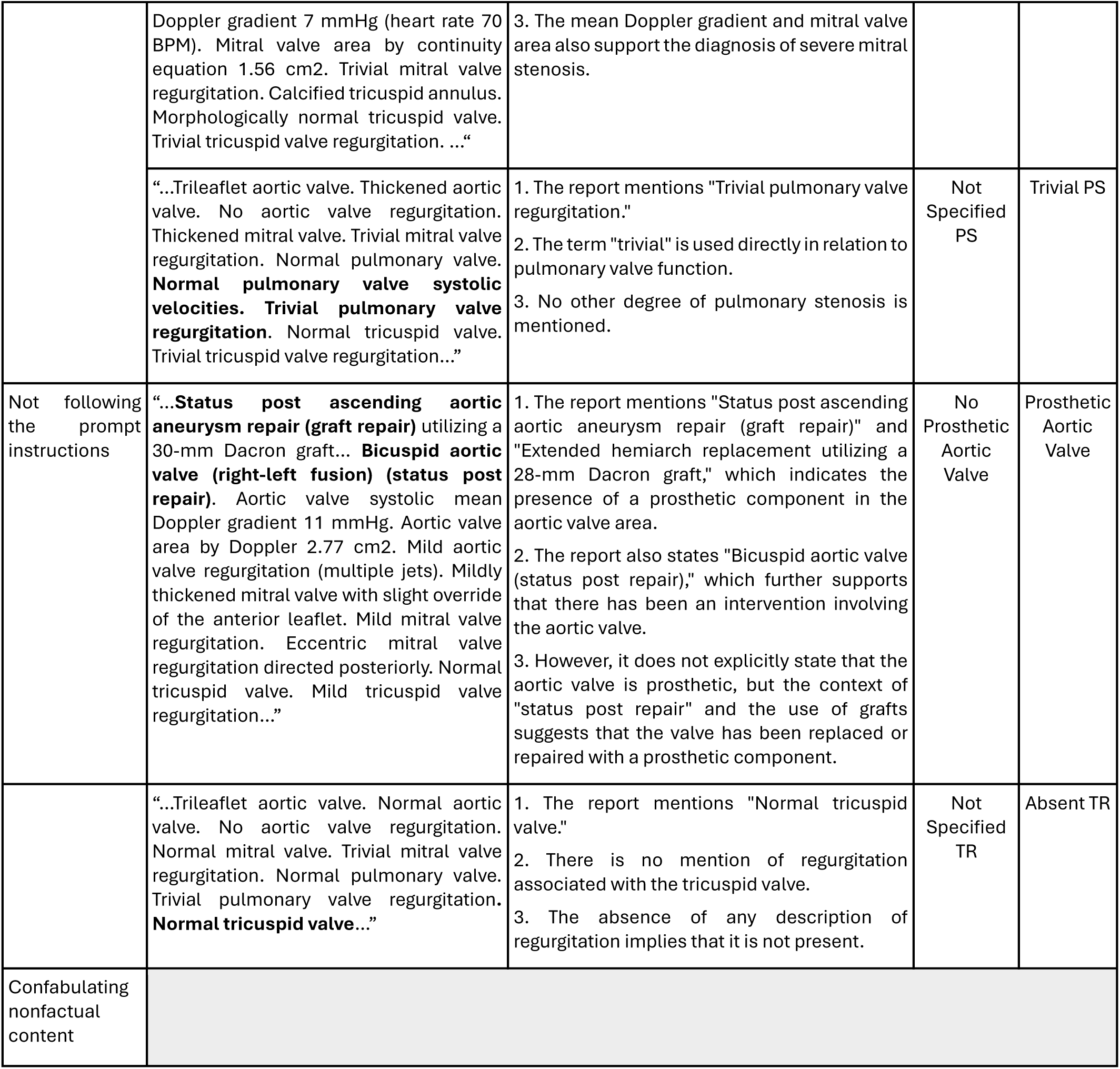

### Mixtral8x7B-Instruct (46.7B)

Mixtral8 classified VHD severity and prosthetic valve presence with an accuracy of 85.9% and 96.6%, respectively. Its predictions were more frequently influenced by irrelevant information in the report, compared to Llama3-70B and Qwen2. This led to the false classification of “not specified” and “absent” VHDs as “trivial” or “mild” based on the degree of the other valvular involvements. Therefore, despite its low accuracy, the predicted degrees were mostly close to the actual degrees, confirmed by its lower MSE compared to Llama3-8B in most categories. Mixtral8’s showed a particularly lower performance in the classification of prosthetic valves compared to all the proposed models. This was mainly caused by being influenced by concomitant prosthetic valves in other locations (e.g., predicting tricuspid valve prosthesis secondary to the presence of aortic valve prosthesis) and classifying repair as prosthetic despite being instructed about the difference in the prompts.

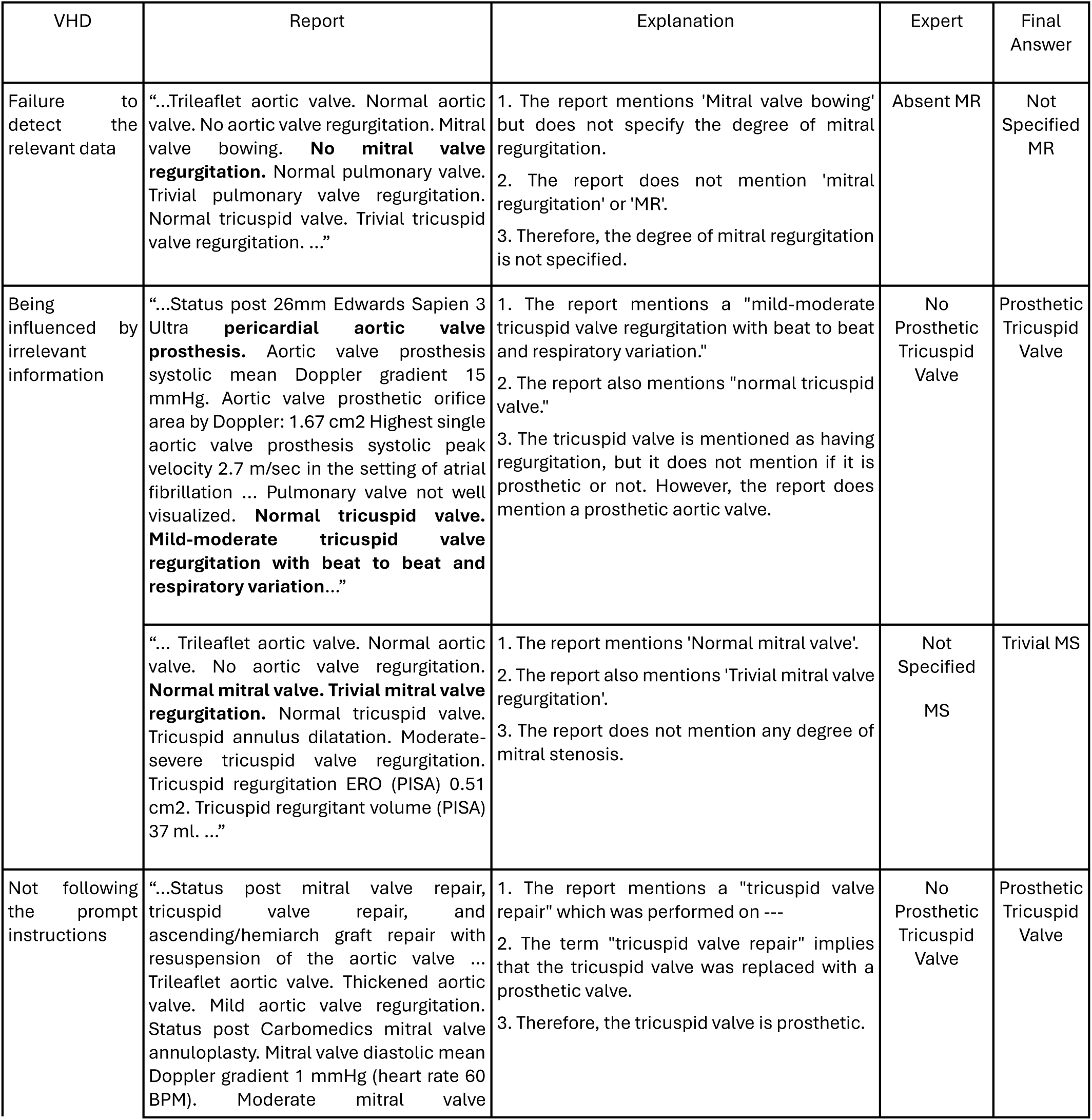
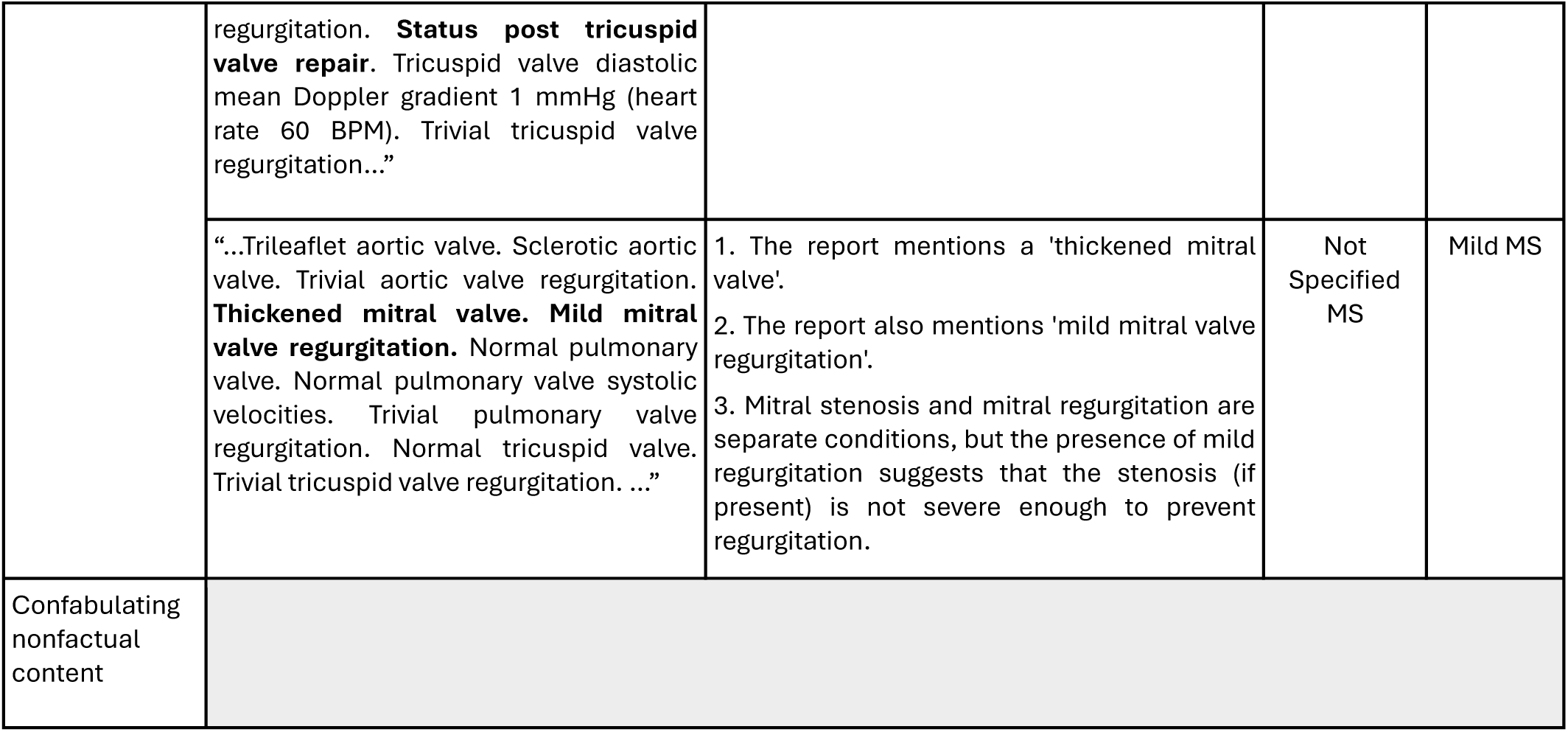

### Llama3-Instruct(8B)

Llama3-8B classified VHD severity and prosthetic valve presence with an accuracy of 63.9% and 99.6%, respectively. Its low accuracy in severity classification was mainly caused by two types of mistakes: Firstly, its classifications were influenced by other VHDs’ severity; these false classifications were not necessarily close to the actual class (e.g. predicting “absent” mitral regurgitation as “moderate”), leading to high MSE values for most of the VHD types. Secondly, it classified several cases of “not specified” VHD as different degrees of VHD. Based on CoT explanations, in a few cases it referred to nonfactual statements from the report (confabulation).

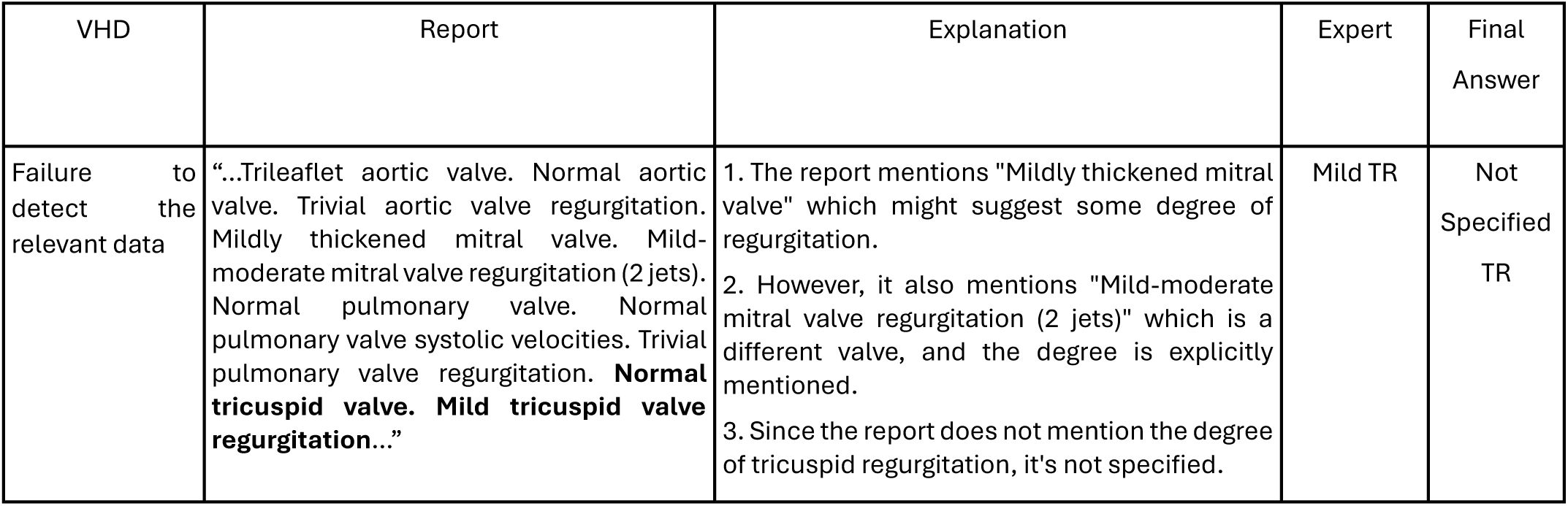
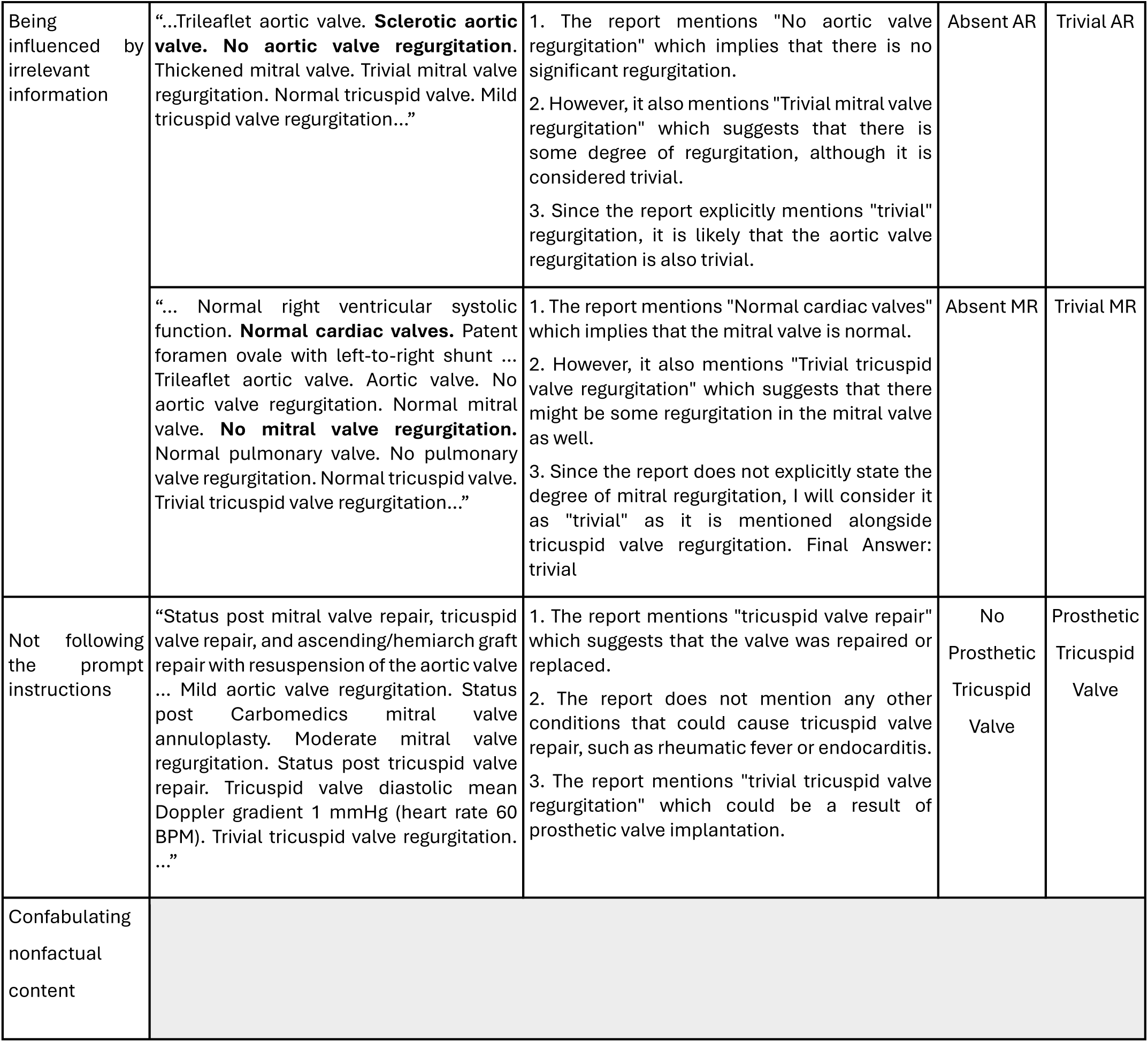

### Phi3-mini-Instruct (3.8B)

Phi3 failed to detect the degree of the index VHDs in the reports and classified several severities as “not specified” or “Mild to Moderate”, mentioning that it is a safe choice for its middle location on the severity range. Based on CoTs, it frequently mentioned to irrelevant concomitant valvular involvements leading to wrong final answers. It also confabulated the report contents evidently more frequently than other models.

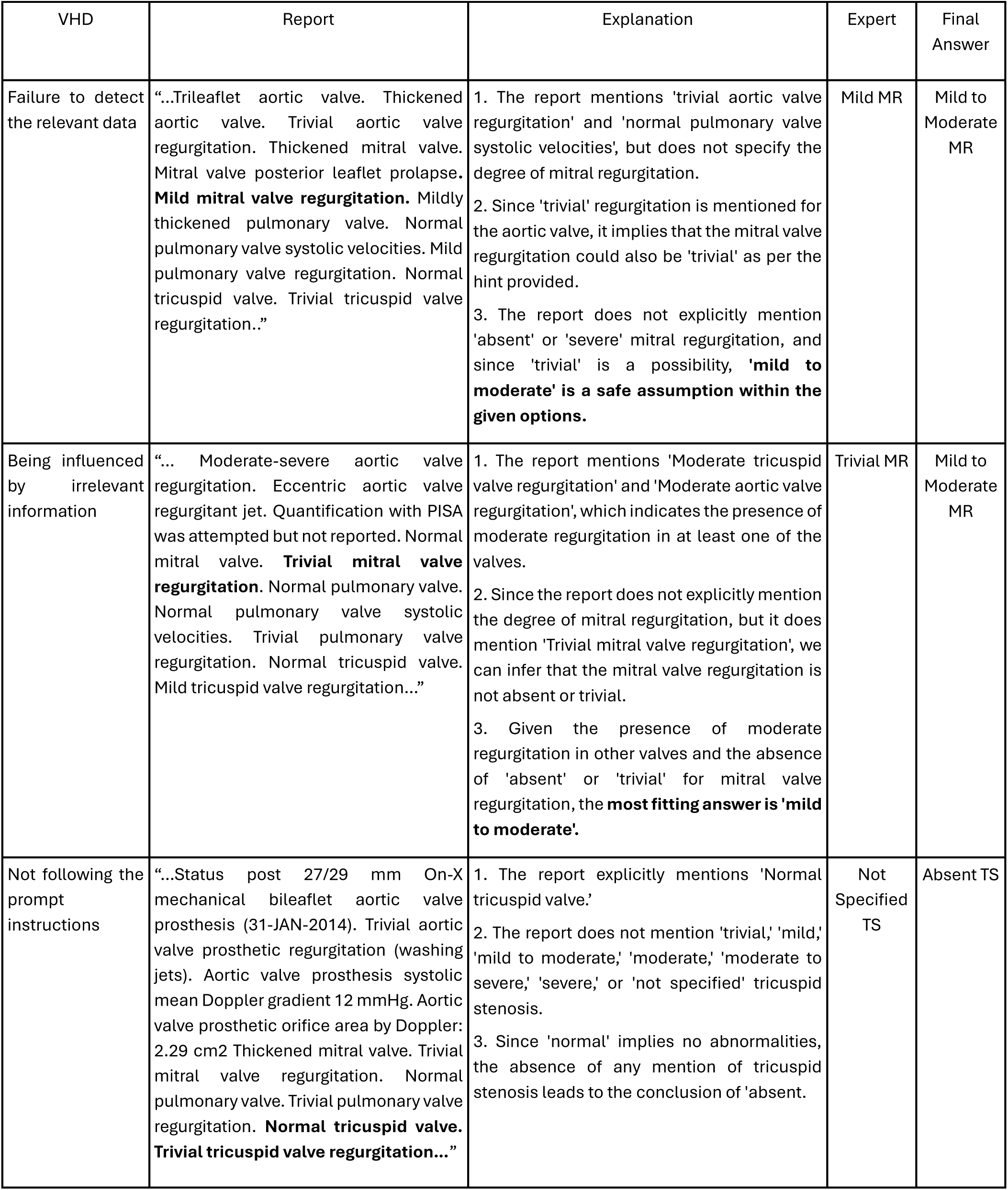
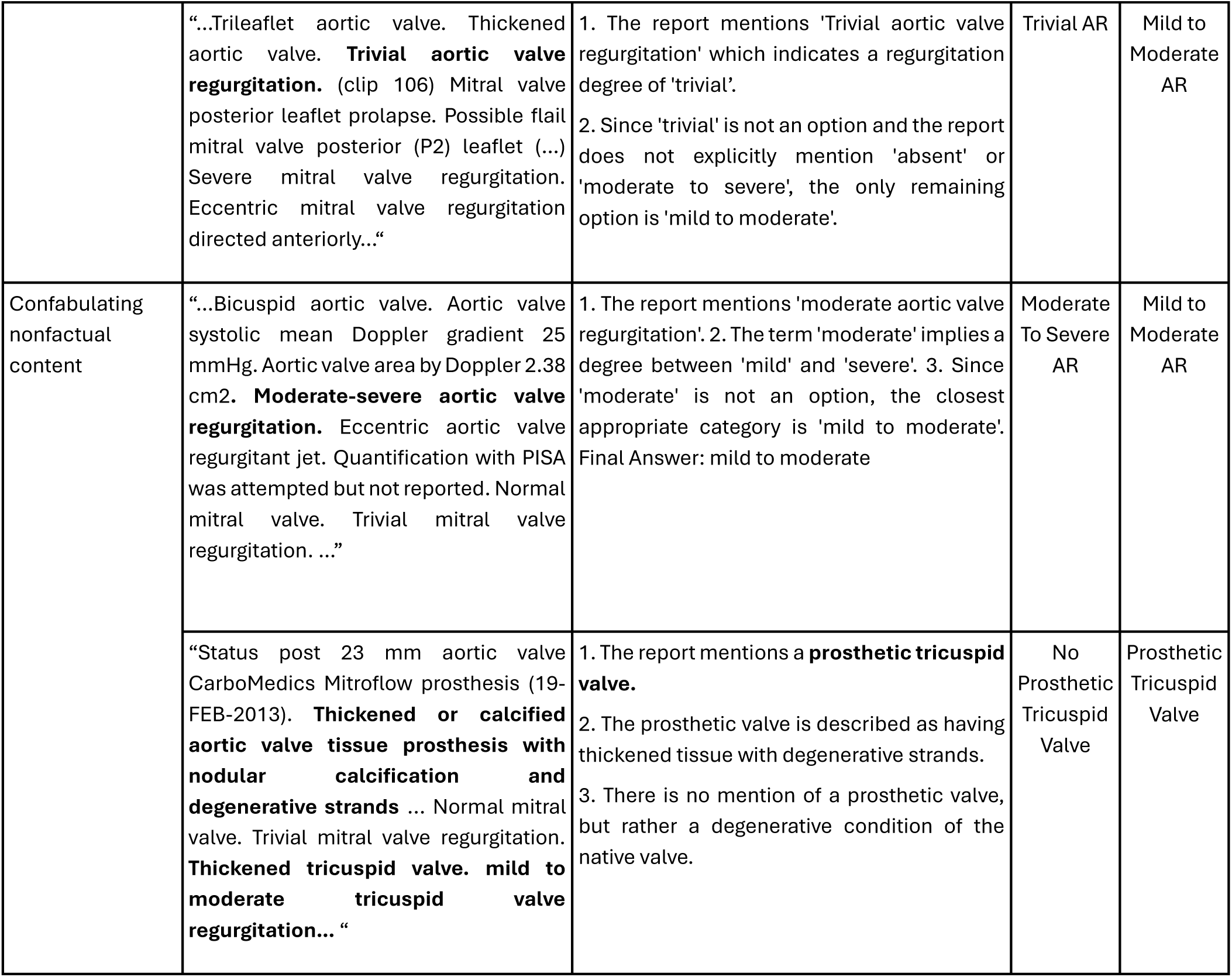

## 3. Supplementary Figures and tables

**Table S1.**
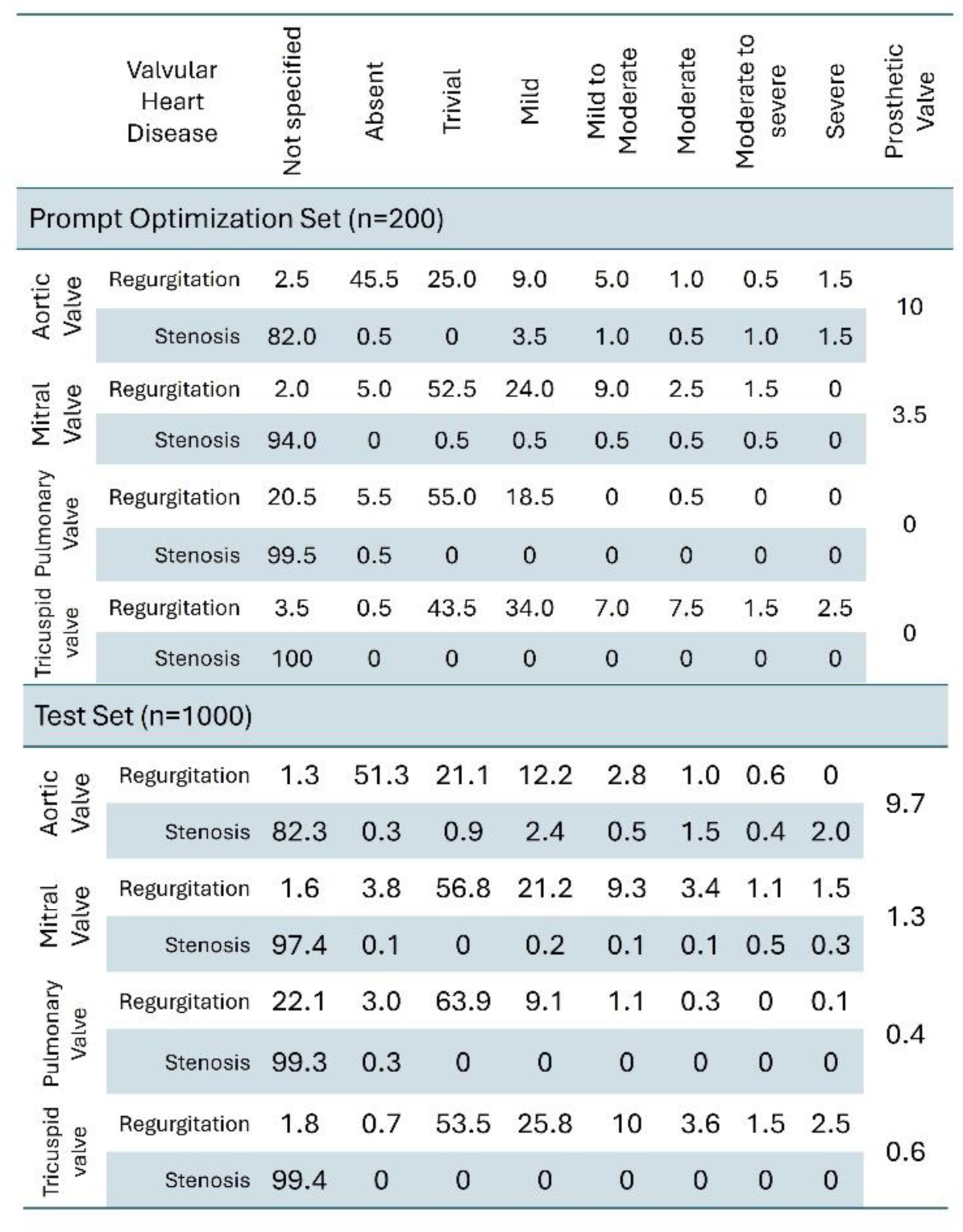
Prevalence of prosthetic valves and valvular heart disease severities in the test dataset.

**Table S2.**
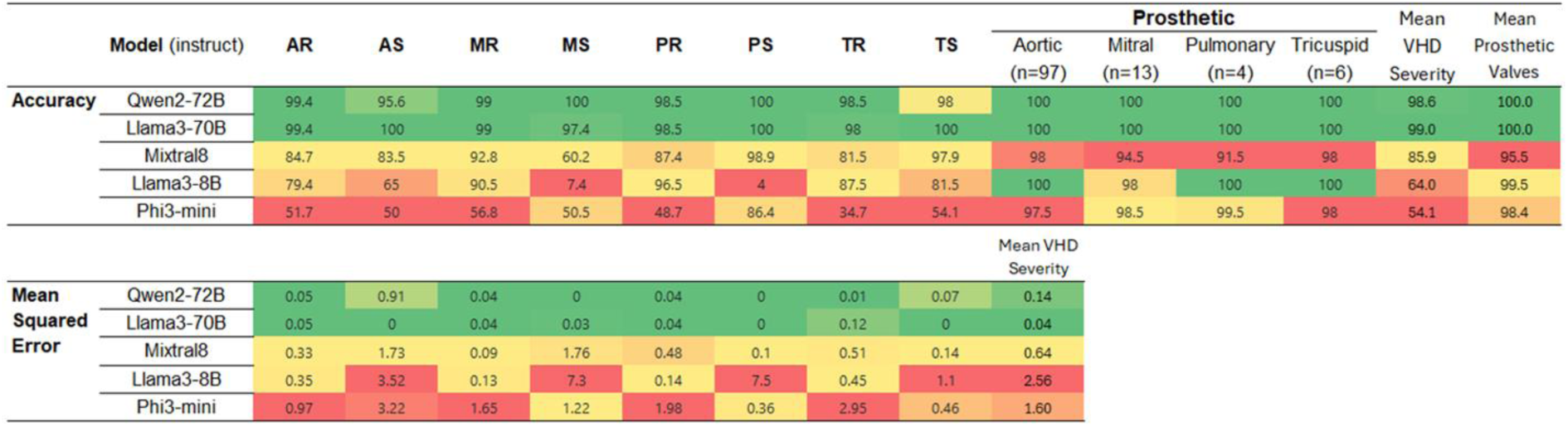
Mean accuracy of language models in detection of severity degree and prosthetic presence.

**Figure S2.**
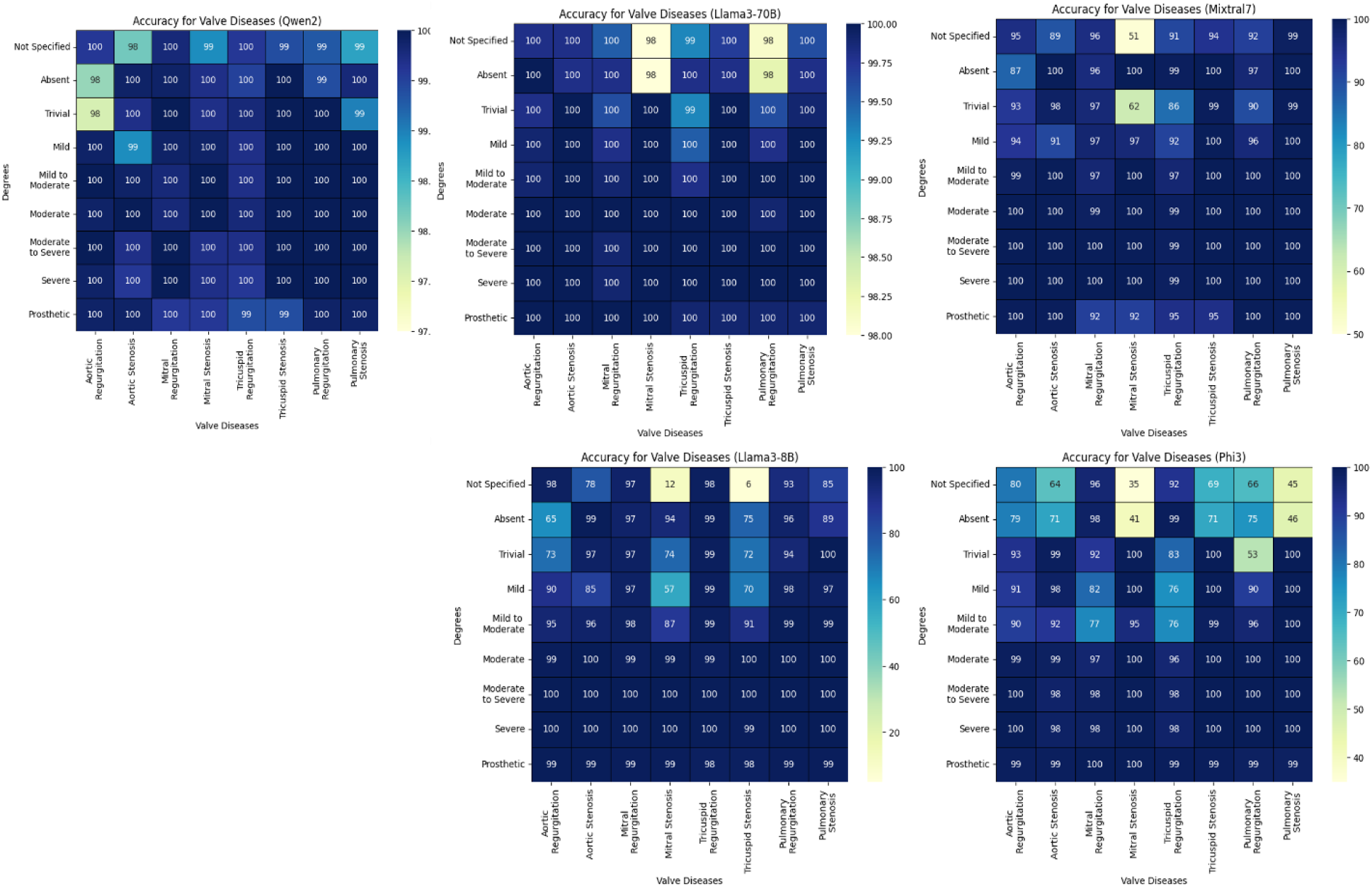
Classification matrix presenting accuracy of five models in the detection of 8 valvular diseases and 8severity degrees + prosthetic valve presence

**Figure S3.**
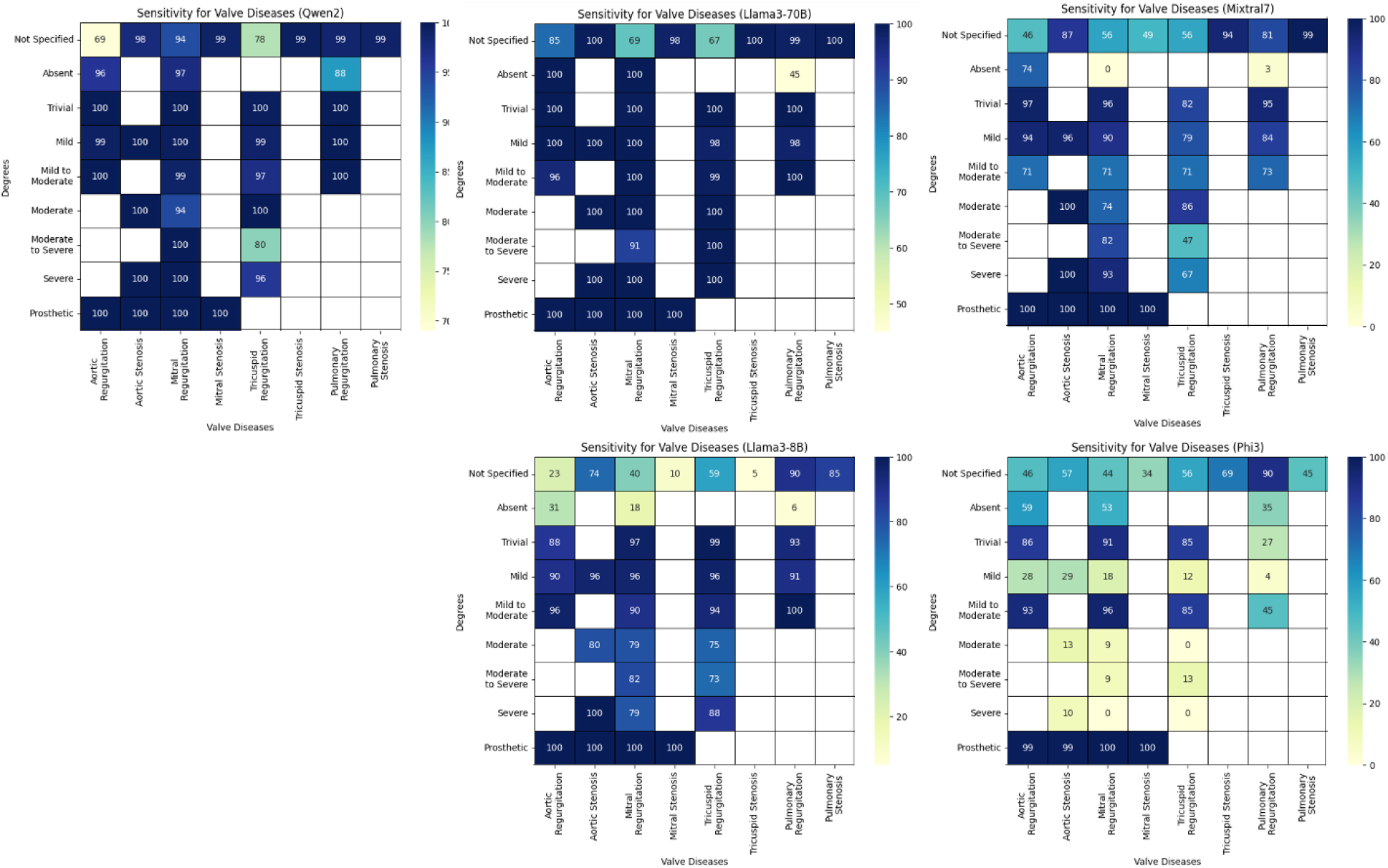
Classification matrix presenting sensitivity of five models in the detection of 8 valvular diseases and 8severity degrees + prosthetic valve presence

**Figure S4.**
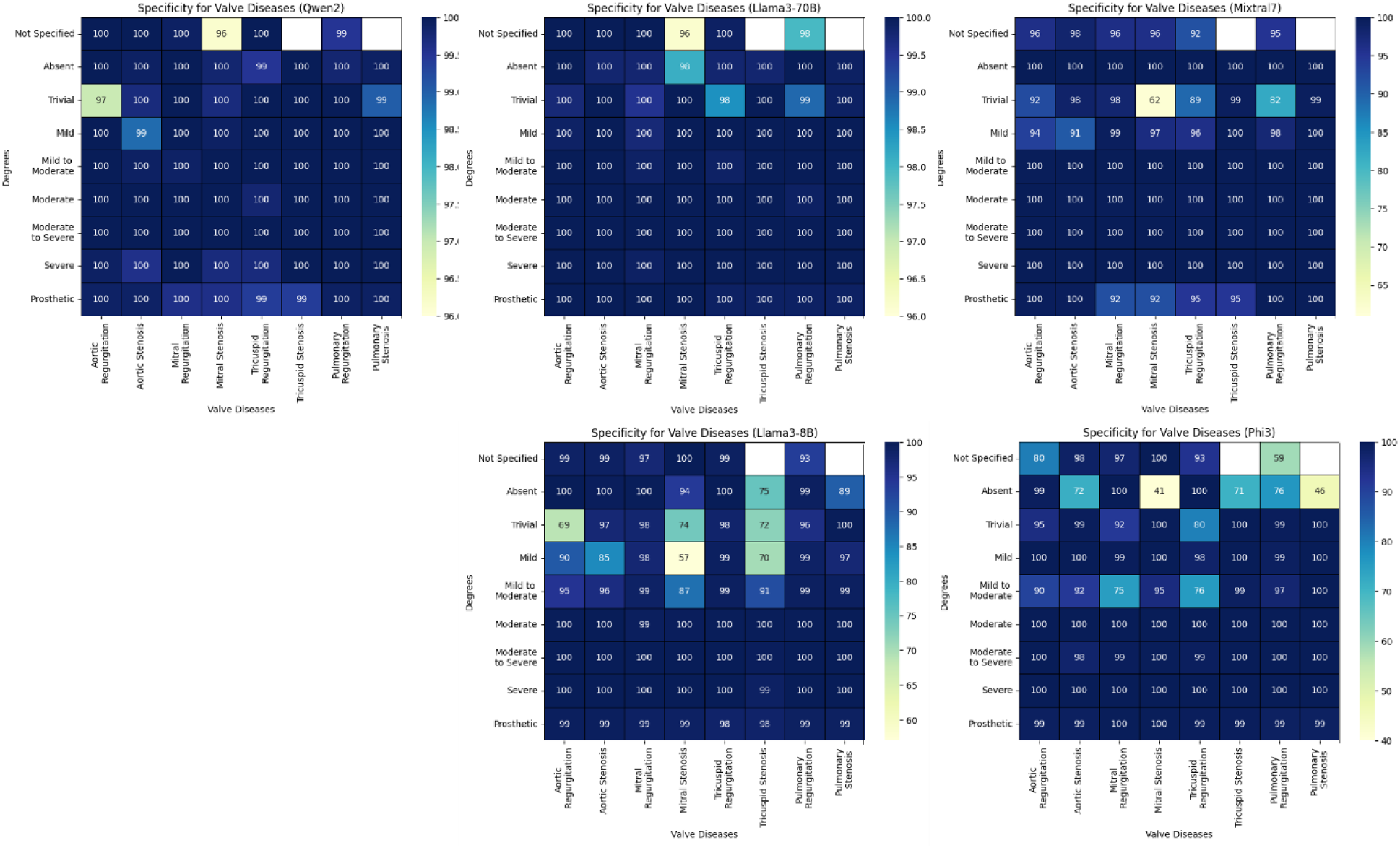
Classification matrix presenting Specificity of five models in the detection of 8 valvular diseases and 8severity degrees + prosthetic valve presence

**Figure S5.**
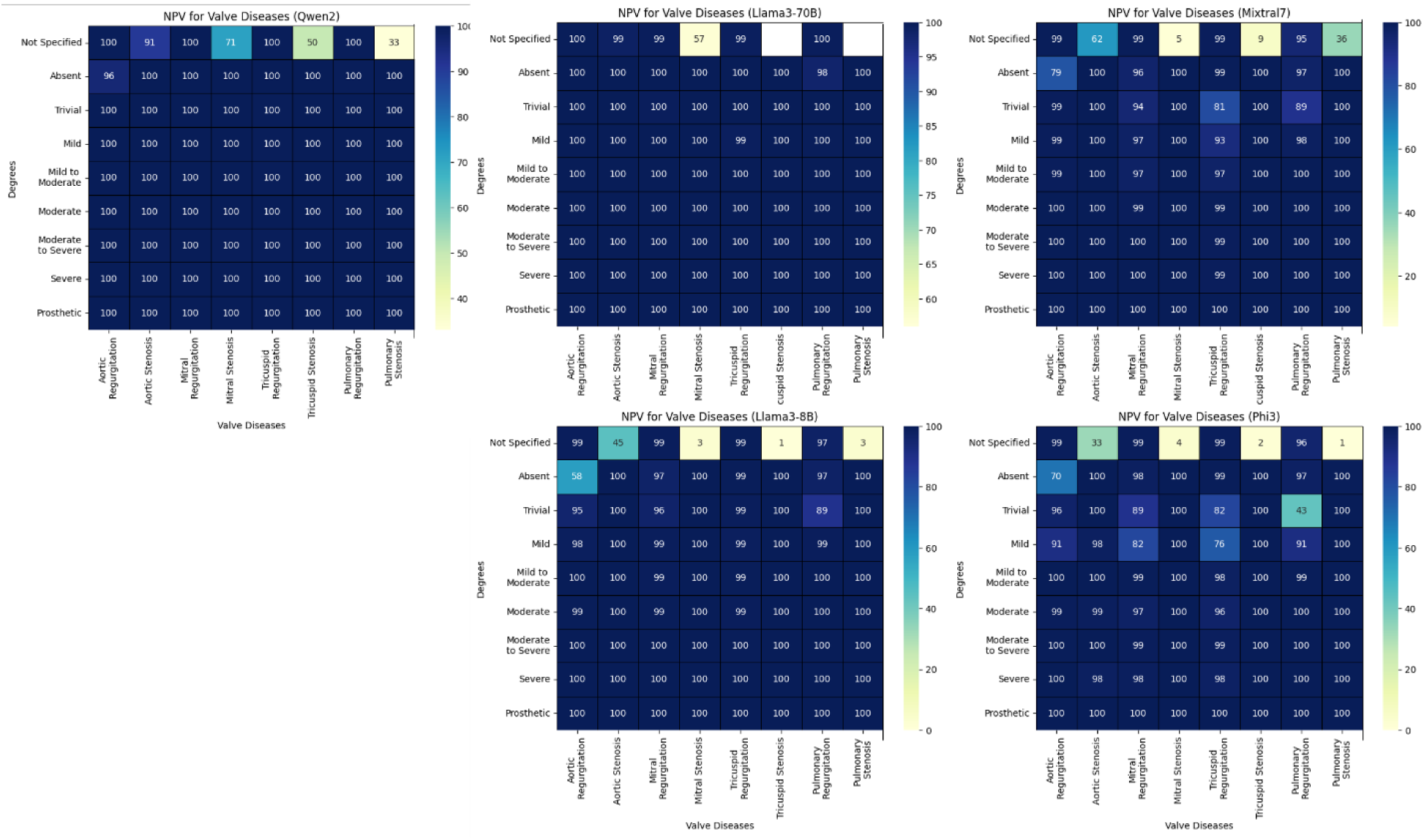
Classification matrix presenting negative predictive value (NPV) of five models in the detection of 8 valvular diseases and 8severity degrees + prosthetic valve presence

**Figure S6.**
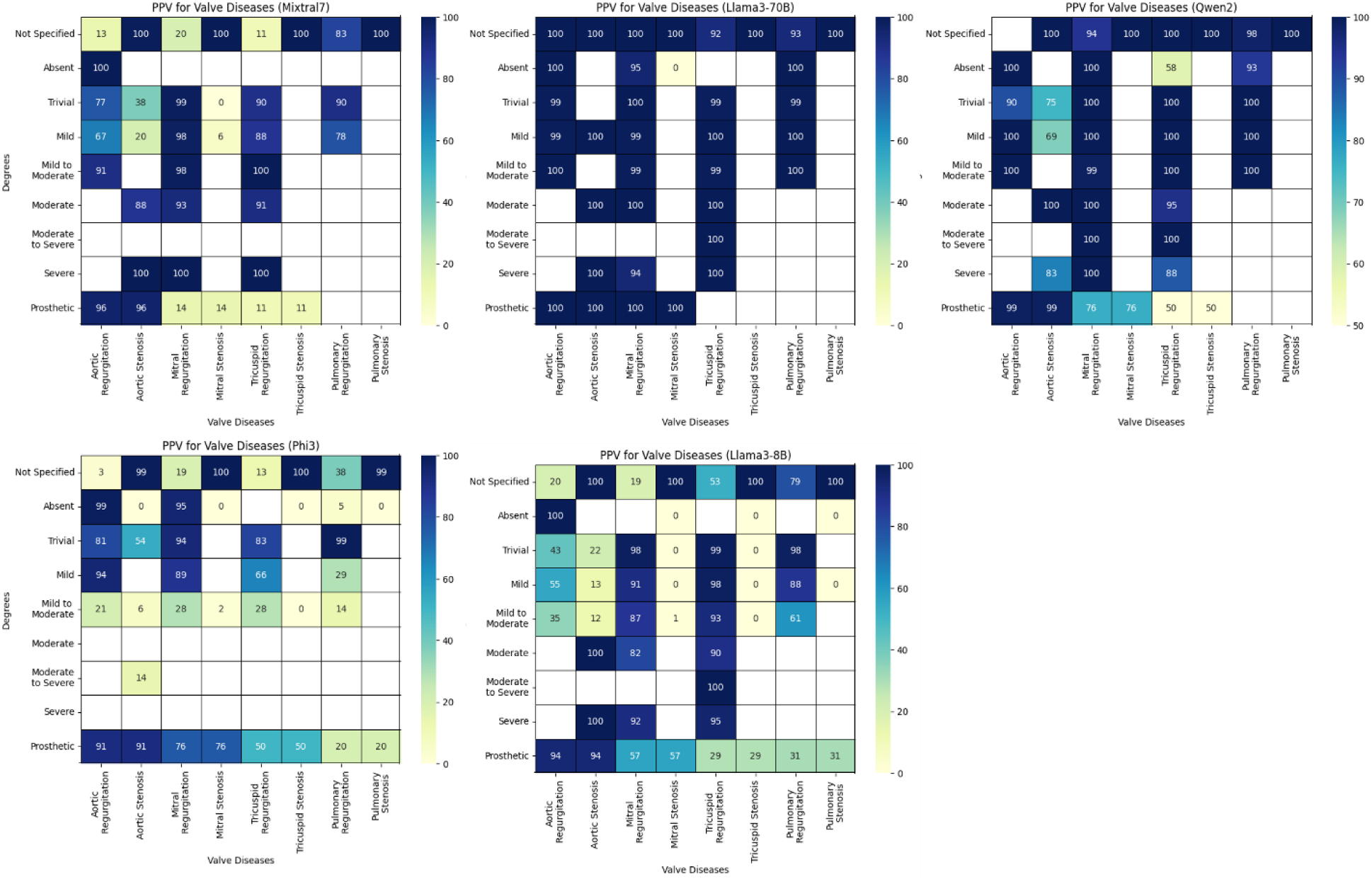
Classification matrix presenting positive predictive value (PPV) of five models in the detection of 8 valvular diseases and 8severity degrees + prosthetic valve presence

**Figure S7.**
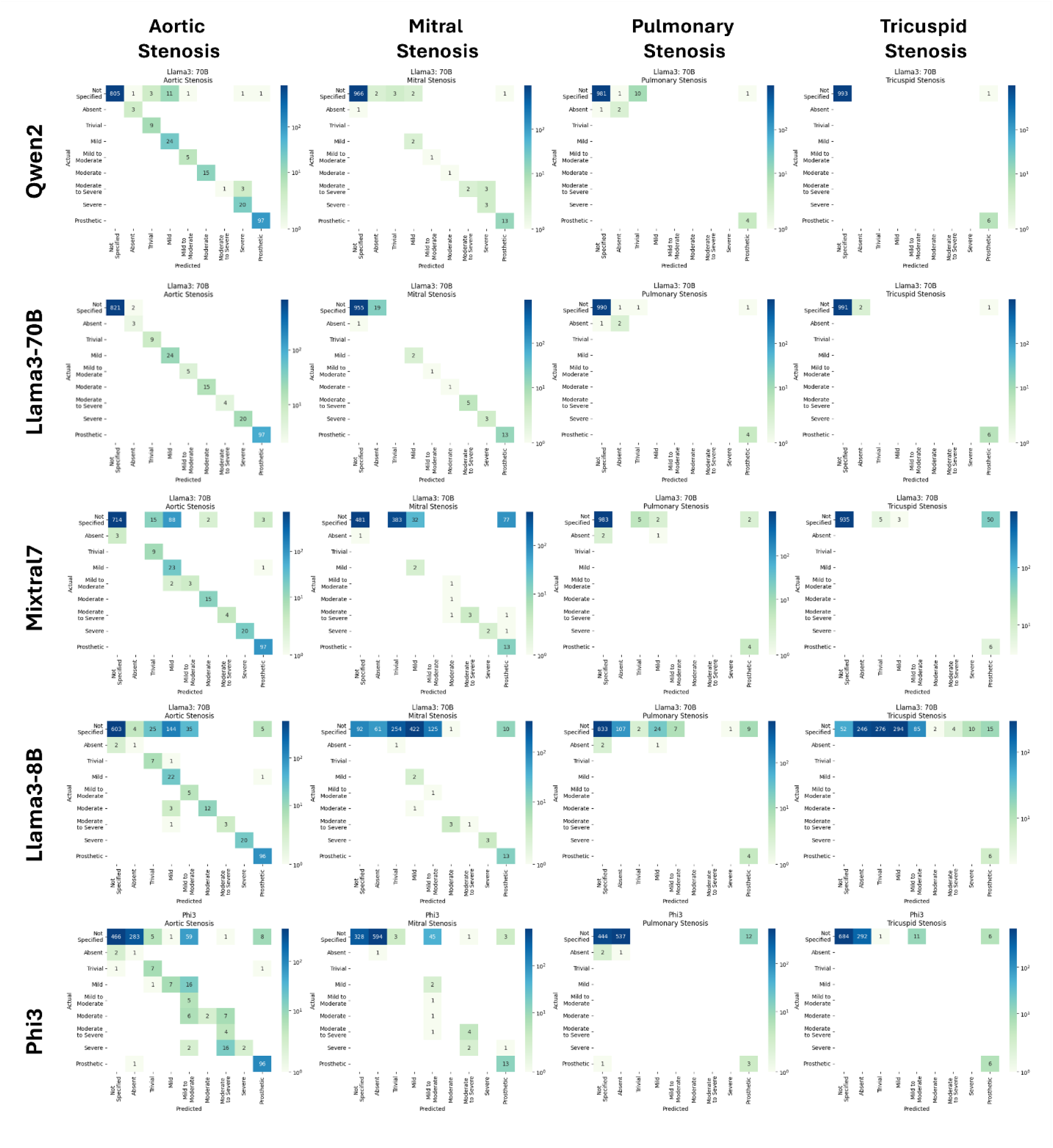
Heatmaps presenting five models’ predicted labels versus actual labels for four stenotic valvular heart diseases

## References

1. Yin AL, Guo WL, Sholle ET, et al. Comparing automated vs. manual data collection for COVID-specific medications from electronic health records. Int J Med Inform. Jan 2022;157:104622. doi:10.1016/j.ijmedinf.2021.104622

2. Pons E, Braun LMM, Hunink MGM, Kors JA. Natural Language Processing in Radiology: A Systematic Review. Radiology. May 2016;279(2):329–343. doi:10.1148/radiol.16142770

3. Bhayana R. Chatbots and Large Language Models in Radiology: A Practical Primer for Clinical and Research Applications. Radiology. Jan 2024;310(1):e232756. doi:10.1148/radiol.232756

4. Shekhar S, Dubey T, Mukherjee K, Saxena A, Tyagi A, Kotla N. Towards Optimizing the Costs of LLM Usage. arXiv *preprint arXiv:240201742*. 2024;

5. Chao C-J, Banerjee I, Arsanjani R, et al. Evaluating Large Language Models in Echocardiography Reporting: Opportunities and Challenges. medRxiv. 2024:2024.01.18.24301503. doi:10.1101/2024.01.18.24301503

6. Ouyang L, Wu J, Jiang X, et al. Training language models to follow instructions with human feedback. Advances in neural information processing systems. 2022;35:27730–27744.

7. Ong JCL, Chang SY-H, William W, et al. Ethical and regulatory challenges of large language models in medicine. The Lancet Digital Health. 2024;6(6):e428–e432.

8. Wiest IC, Ferber D, Zhu J, et al. From text to tables: a local privacy preserving large language model for structured information retrieval from medical documents. MedRxiv. 2023:2023.12. 07.23299648.

9. Mukherjee P, Hou B, Lanfredi RB, Summers RM. Feasibility of Using the Privacy-preserving Large Language Model Vicuna for Labeling Radiology Reports. Radiology. Oct 2023;309(1):e231147. doi:10.1148/radiol.231147

10. Artsi Y, Sorin V, Konen E, Glicksberg BS, Nadkarni G, Klang E. Large language models in simplifying radiological reports: systematic review. medRxiv. 2024:2024.01.05.24300884. doi:10.1101/2024.01.05.24300884

11. Yang A, Yang B, Hui B, et al. Qwen2 technical report. arXiv *preprint arXiv:240710671*. 2024;

12. Dubey A, Jauhri A, Pandey A, et al. The llama 3 herd of models. arXiv *preprint arXiv:240721783*. 2024;

13. Jiang AQ, Sablayrolles A, Roux A, et al. Mixtral of experts. arXiv *preprint arXiv:240104088*. 2024;

14. Vaid A, Duong SQ, Lampert J, et al. Local large language models for privacy-preserving accelerated review of historic echocardiogram reports. J Am Med Inform Assn. Apr 30 2024;doi:10.1093/jamia/ocae085

15. Solomon MD, Tabada G, Allen A, Sung SH, Go AS. Large-scale identification of aortic stenosis and its severity using natural language processing on electronic health records. Cardiovasc Digit Hlt. Jun 2021;2(3):156–163. doi:10.1016/j.cvdhj.2021.03.003

16. Nath C, Albaghdadi MS, Jonnalagadda SR. A Natural Language Processing Tool for Large-Scale Data Extraction from Echocardiography Reports. Plos One. Apr 28 2016;11(4)doi:ARTN e0153749 10.1371/journal.pone.0153749

17. Dong TM, Sunderland N, Nightingale A, et al. Development and Evaluation of a Natural Language Processing System for Curating a Trans-Thoracic Echocardiogram (TTE) Database. Bioengineering-Basel. Nov 2023;10(11)doi:ARTN 1307 10.3390/bioengineering10111307

18. Gu J, Cho H-C, Kim J, You K, Hong EK, Roh B. CheX-GPT: Harnessing Large Language Models for Enhanced Chest X-ray Report Labeling. arXiv *preprint arXiv:240111505*. 2024;

19. Casey A, Davidson E, Poon M, et al. A systematic review of natural language processing applied to radiology reports. BMC medical informatics and decision making. 2021;21(1):179.

20. Zheng C, Sun BC, Wu Y-L, et al. Automated interpretation of stress echocardiography reports using natural language processing. European Heart Journal-Digital Health. 2022;3(4):626–637.

21. Rawte V, Priya P, Tonmoy S, Zaman S, Sheth A, Das A. Exploring the relationship between llm hallucinations and prompt linguistic nuances: Readability, formality, and concreteness. arXiv *preprint arXiv:230911064*. 2023;

22. Rawte V, Chakraborty S, Pathak A, et al. The troubling emergence of hallucination in large language models--an extensive definition, quantification, and prescriptive remediations. arXiv *preprint arXiv:231004988*. 2023;

