## Supplementary methods and results for "A Comparative Analysis of Privacy-Preserving Large Language Models For Automated Echocardiography Report Analysis"

| VHD | Report | Explanation | Expert | Final answer |
|---|---|---|---|---|
| Failure to detect the relevant data | | | | |
| Being influenced by irrelevant information | "…Trileaflet aortic valve. Normal aortic valve. No aortic valve regurgitation. Normal mitral valve. Trivial mitral valve regurgitation. Normal tricuspid valve. Trivial tricuspid valve regurgitation. **Normal pulmonary valve. Trivial pulmonary valve regurgitation.** OTHER ECHO FINDINGS: Normal inferior vena cava size with reduced inspiratory collapse. **Trivial left pulmonary artery stenosis.** Normal scan of the aorta…" | 1. The report mentions "Trivial left pulmonary artery stenosis" which indicates the presence of pulmonary stenosis, but to a trivial degree.<br>2. There is no mention of other degrees of pulmonary stenosis, such as mild, moderate, or severe.<br>3. The report explicitly states "trivial" pulmonary stenosis, which means it is not absent or not specified. | Not Specified PS | Trivial PS |
| Not following the prompt instructions | "…Trileaflet aortic valve. Normal aortic valve. No aortic valve regurgitation. **Normal mitral valve. Trivial mitral valve regurgitation.** Normal pulmonary valve. Trivial pulmonary valve regurgitation. Normal tricuspid valve. Trivial tricuspid valve regurgitation…" | 1. The report mentions "Normal mitral valve" which implies that there is no mitral stenosis.<br>2. There is no explicit mention of mitral stenosis or its degree in the report.<br>3. The report does not mention "no mitral stenosis" explicitly, but the normalcy of the mitral valve suggests its absence. | Not Specified MS | Absent MS |
| | "…Normal pulmonary valve. Trivial pulmonary valve regurgitation**. Status post Carbomedics tricuspid valve annuloplasty.** Tricuspid valve diastolic mean Doppler gradient 4 mmHg (heart rate 92 BPM). Mild-moderate tricuspid valve regurgitation…" | 1. The report mentions "Status post Carbomedics tricuspid valve annuloplasty" which suggests a prosthetic tricuspid valve.<br>2. The term "annuloplasty" implies a surgical repair or replacement of the valve. | No Prosthetic Tricuspid Valve | Prosthetic Tricuspid Valve |
| Confabulating nonfactual content | | | | |

**Qwen2-Instruct (72.7B):**

Qwen2 showed the second-best performance with an accuracy of 98.9% in the classification of VHD severities and 99.9% in the classification of prosthetic valve presence. Its most common mistake was to be influenced by irrelevant information in the report, rather than classifying the VHD degree based on the explicit degree mentioned. An example was classifying the sclerotic or thickened aortic valves as "mild AS", while the degree of stenosis was not explicitly mentioned throughout the report and this concept was particularly clarified for the LLM in the prompts. This led to an accuracy of 97% in the classification of AR and AS. Qwen2 had the same problem of classifying moderate to severe as severe in a few cases and false classification of an aortic, a mitral, and a pulmonary repair as prosthetic valve.

| VHD | Report | Explanation | Expert | Final Answer |
|---|---|---|---|---|
| Failure to detect the relevant data | | | | |
| Being influenced by irrelevant information | "...Trileaflet aortic valve. Sclerotic aortic valve. No aortic valve regurgitation. **Moderate-severe calcific mitral valve stenosis. Severely calcified mitral annulus**. Mitral valve diastolic mean Doppler gradient 7 mmHg (heart rate 70 BPM). Mitral valve area by continuity equation 1.56 cm2. Trivial mitral valve regurgitation. Calcified tricuspid annulus. Morphologically normal tricuspid valve. Trivial tricuspid valve regurgitation. ..." | 1. The report specifically mentions "Moderate-severe calcific mitral valve stenosis."<br>2. It further details the severity with "Severely calcified mitral annulus."<br>3. The mean Doppler gradient and mitral valve area also support the diagnosis of severe mitral stenosis. | Moderate to Severe MS | Severe MS |
| | "...Trileaflet aortic valve. Thickened aortic valve. No aortic valve regurgitation. Thickened mitral valve. Trivial mitral valve regurgitation. Normal pulmonary valve. **Normal pulmonary valve systolic velocities. Trivial pulmonary valve regurgitation**. Normal tricuspid valve. Trivial tricuspid valve regurgitation..." | 1. The report mentions "Trivial pulmonary valve regurgitation."<br>2. The term "trivial" is used directly in relation to pulmonary valve function.<br>3. No other degree of pulmonary stenosis is mentioned. | Not Specified PS | Trivial PS |
| Not following the prompt instructions | "...**Status post ascending aortic aneurysm repair (graft repair)** utilizing a [...] graft... **Bicuspid aortic valve (right-left fusion) (status post repair)**. Aortic valve systolic mean Doppler gradient 11 mmHg. Aortic valve area by Doppler 2.77 cm2. Mild aortic valve regurgitation (multiple jets). Mildly thickened mitral valve with slight override of the anterior leaflet. Mild mitral valve regurgitation. Eccentric mitral valve regurgitation directed posteriorly. Normal tricuspid valve. Mild tricuspid valve regurgitation..." | 1. The report mentions "Status post ascending aortic aneurysm repair (graft repair)" and "Extended hemiarch replacement utilizing a 28-mm Dacron graft," which indicates the presence of a prosthetic component in the aortic valve area.<br>2. The report also states "Bicuspid aortic valve (status post repair)," which further supports that there has been an intervention involving the aortic valve.<br>3. However, it does not explicitly state that the aortic valve is prosthetic, but the context of "status post repair" and the use of grafts suggests that the valve has been replaced or repaired with a prosthetic component. | No Prosthetic Aortic Valve | Prosthetic Aortic Valve |
| | "...Trileaflet aortic valve. Normal aortic valve. No aortic valve regurgitation. Normal mitral valve. Trivial mitral valve regurgitation. Normal pulmonary valve. | 1. The report mentions "Normal tricuspid valve."<br>2. There is no mention of regurgitation associated with the tricuspid valve. | Not Specified TR | Absent TR |

| | | |
|---|---|---|
| | Trivial pulmonary valve regurgitation. **Normal tricuspid valve**…” | 3. The absence of any description of regurgitation implies that it is not present. |
| Confabulating nonfactual content | | |

**Mixtral8x7B-Instruct (46.7B):**

Mixtral8 classified VHD severity and prosthetic valve presence with an accuracy of 85.9% and 96.6%, respectively. Its predictions were more frequently influenced by irrelevant information in the report, compared to Llama3-70B and Qwen2. This led to the false classification of “not specified” and “absent” VHDs as “trivial” or “mild” based on the degree of the other valvular involvements. Therefore, despite its low accuracy, the predicted degrees were mostly close to the actual degrees, confirmed by its lower MSE compared to Llama3-8B in most categories. Mixtral8’s showed a particularly lower performance in the classification of prosthetic valves compared to all the proposed models. This was mainly caused by being influenced by concomitant prosthetic valves in other locations (e.g., predicting tricuspid valve prosthesis secondary to the presence of aortic valve prosthesis) and classifying repair as prosthetic despite being instructed about the difference in the prompts.

| VHD | Report | Explanation | Expert | Final Answer |
|---|---|---|---|---|
| Failure to detect the relevant data | “…Trileaflet aortic valve. Normal aortic valve. No aortic valve regurgitation. Mitral valve bowing. **No mitral valve regurgitation.** Normal pulmonary valve. Trivial pulmonary valve regurgitation. Normal tricuspid valve. Trivial tricuspid valve regurgitation. …” | 1. The report mentions 'Mitral valve bowing' but does not specify the degree of mitral regurgitation.<br>2. The report does not mention 'mitral regurgitation' or 'MR'.<br>3. Therefore, the degree of mitral regurgitation is not specified. | Absent MR | Not Specified MR |

| | | | | |
|---|---|---|---|---|
| Being influenced by irrelevant information | "...Status post [...] **pericardial aortic valve prosthesis.** Aortic valve prosthesis systolic mean Doppler gradient 15 mmHg. Aortic valve prosthetic orifice area by Doppler: 1.67 cm2 Highest single aortic valve prosthesis systolic peak velocity 2.7 m/sec in the setting of atrial fibrillation ... Pulmonary valve not well visualized. **Normal tricuspid valve. Mild-moderate tricuspid valve regurgitation with beat to beat and respiratory variation**..." | 1. The report mentions a "mild-moderate tricuspid valve regurgitation with beat to beat and respiratory variation."<br>2. The report also mentions "normal tricuspid valve."<br>3. The tricuspid valve is mentioned as having regurgitation, but it does not mention if it is prosthetic or not. However, the report does mention a prosthetic aortic valve. | No Prosthetic Tricuspid Valve | Prosthetic Tricuspid Valve |
| | "... Trileaflet aortic valve. Normal aortic valve. No aortic valve regurgitation. **Normal mitral valve. Trivial mitral valve regurgitation.** Normal tricuspid valve. Tricuspid annulus dilatation. Moderate-severe tricuspid valve regurgitation. Tricuspid regurgitation ERO (PISA) 0.51 cm2. Tricuspid regurgitant volume (PISA) 37 ml. ..." | 1. The report mentions 'Normal mitral valve'.<br>2. The report also mentions 'Trivial mitral valve regurgitation'.<br>3. The report does not mention any degree of mitral stenosis. | Not Specified<br><br>MS | Trivial MS |
| Not following the prompt instructions | "...Status post mitral valve repair, tricuspid valve repair, and ascending/hemiarch graft repair with resuspension of the aortic valve ... Trileaflet aortic valve. Thickened aortic valve. Mild aortic valve regurgitation. Status post Carbomedics mitral valve annuloplasty. Mitral valve diastolic mean Doppler gradient 1 mmHg (heart rate 60 BPM). Moderate mitral valve regurgitation. **Status post tricuspid valve repair**. Tricuspid valve diastolic mean Doppler gradient 1 mmHg (heart rate 60 BPM). Trivial tricuspid valve regurgitation..." | 1. The report mentions a "tricuspid valve repair" which was performed on [...]<br>2. The term "tricuspid valve repair" implies that the tricuspid valve was replaced with a prosthetic valve.<br>3. Therefore, the tricuspid valve is prosthetic. | No Prosthetic Tricuspid Valve | Prosthetic Tricuspid Valve |
| | "...Trileaflet aortic valve. Sclerotic aortic valve. Trivial aortic valve regurgitation. **Thickened mitral valve. Mild mitral valve regurgitation.** Normal pulmonary valve. Normal pulmonary valve systolic velocities. Trivial pulmonary valve regurgitation. Normal tricuspid valve. Trivial tricuspid valve regurgitation. ..." | 1. The report mentions a 'thickened mitral valve'.<br>2. The report also mentions 'mild mitral valve regurgitation'.<br>3. Mitral stenosis and mitral regurgitation are separate conditions, but the presence of mild regurgitation suggests that the stenosis (if present) is not severe enough to prevent regurgitation. | Not Specified MS | Mild MS |
| Confabulating nonfactual content | | | | |

### Llama3-Instruct(8B):

Llama3-8B classified VHD severity and prosthetic valve presence with an accuracy of 63.9% and 99.6%, respectively. Its low accuracy in severity classification was mainly caused by two types of mistakes: Firstly, its classifications were influenced by other VHDs' severity; these false classifications were not necessarily close to the actual class (e.g. predicting "absent" mitral regurgitation as "moderate"), leading to high MSE values for most of the VHD types. Secondly, it classified several cases of "not specified" VHD as different degrees of VHD. Based on CoT explanations, in a few cases it referred to nonfactual statements from the report (confabulation).

| VHD | Report | Explanation | Expert | Final Answer |
|---|---|---|---|---|
| Failure to detect the relevant data | "...Trileaflet aortic valve. Normal aortic valve. Trivial aortic valve regurgitation. Mildly thickened mitral valve. Mild-moderate mitral valve regurgitation (2 jets). Normal pulmonary valve. Normal pulmonary valve systolic velocities. Trivial pulmonary valve regurgitation. **Normal tricuspid valve. Mild tricuspid valve regurgitation**..." | 1. The report mentions "Mildly thickened mitral valve" which might suggest some degree of regurgitation.<br>2. However, it also mentions "Mild-moderate mitral valve regurgitation (2 jets)" which is a different valve, and the degree is explicitly mentioned.<br>3. Since the report does not mention the degree of tricuspid regurgitation, it's not specified. | Mild TR | Not Specified TR |
| Being influenced by irrelevant information | "...Trileaflet aortic valve. **Sclerotic aortic valve. No aortic valve regurgitation**. Thickened mitral valve. Trivial mitral valve regurgitation. Normal tricuspid valve. Mild tricuspid valve regurgitation..." | 1. The report mentions "No aortic valve regurgitation" which implies that there is no significant regurgitation.<br>2. However, it also mentions "Trivial mitral valve regurgitation" which suggests that there is some degree of regurgitation, although it is considered trivial.<br>3. Since the report explicitly mentions "trivial" regurgitation, it is likely that the aortic valve regurgitation is also trivial. | Absent AR | Trivial AR |
| | "... Normal right ventricular systolic function. **Normal cardiac valves.** Patent foramen ovale with left-to-right shunt ... Trileaflet aortic valve. Aortic valve. No aortic valve regurgitation. Normal mitral valve. **No mitral valve regurgitation.** Normal pulmonary valve. No pulmonary valve regurgitation. Normal tricuspid valve. Trivial tricuspid valve regurgitation..." | 1. The report mentions "Normal cardiac valves" which implies that the mitral valve is normal.<br>2. However, it also mentions "Trivial tricuspid valve regurgitation" which suggests that there might be some regurgitation in the mitral valve as well.<br>3. Since the report does not explicitly state the degree of mitral regurgitation, I will consider it as "trivial" as it is mentioned alongside tricuspid valve regurgitation. Final Answer: trivial | Absent MR | Trivial MR |

| | | | | |
|---|---|---|---|---|
| Not following the prompt instructions | "Status post mitral valve repair, tricuspid valve repair, and ascending/hemiarch graft repair with resuspension of the aortic valve … Mild aortic valve regurgitation. Status post Carbomedics mitral valve annuloplasty. Moderate mitral valve regurgitation. Status post tricuspid valve repair. Tricuspid valve diastolic mean Doppler gradient 1 mmHg (heart rate 60 BPM). Trivial tricuspid valve regurgitation. …" | 1. The report mentions "tricuspid valve repair" which suggests that the valve was repaired or replaced.<br>2. The report does not mention any other conditions that could cause tricuspid valve repair, such as rheumatic fever or endocarditis.<br>3. The report mentions "trivial tricuspid valve regurgitation" which could be a result of prosthetic valve implantation. | No Prosthetic Tricuspid Valve | Prosthetic Tricuspid Valve |
| Confabulating nonfactual content | | | | |

**Phi3-mini-Instruct (3.8B):**

Phi3 failed to detect the degree of the index VHDs in the reports and classified several severities as "not specified" or "Mild to Moderate", mentioning that it is a safe choice for its middle location on the severity range. Based on CoTs, it frequently mentioned to irrelevant concomitant valvular involvements leading to wrong final answers. It also confabulated the report contents evidently more frequently than other models.

| VHD | Report | Explanation | Expert | Final Answer |
|---|---|---|---|---|
| Failure to detect the relevant data | "…Trileaflet aortic valve. Thickened aortic valve. Trivial aortic valve regurgitation. Thickened mitral valve. Mitral valve posterior leaflet prolapse. **Mild mitral valve regurgitation.** Mildly thickened pulmonary valve. Normal pulmonary valve systolic velocities. Mild pulmonary valve regurgitation. Normal tricuspid valve. Trivial tricuspid valve regurgitation.." | 1. The report mentions 'trivial aortic valve regurgitation' and 'normal pulmonary valve systolic velocities', but does not specify the degree of mitral regurgitation.<br>2. Since 'trivial' regurgitation is mentioned for the aortic valve, it implies that the mitral valve regurgitation could also be 'trivial' as per the hint provided.<br>3. The report does not explicitly mention 'absent' or 'severe' mitral regurgitation, and since 'trivial' is a possibility, **'mild to moderate' is a safe assumption within the given options.** | Mild MR | Mild to Moderate MR |

| | | | | |
|---|---|---|---|---|
| Being influenced by irrelevant information | "... Moderate-severe aortic valve regurgitation. Eccentric aortic valve regurgitant jet. Quantification with PISA was attempted but not reported. Normal mitral valve. **Trivial mitral valve regurgitation**. Normal pulmonary valve. Normal pulmonary valve systolic velocities. Trivial pulmonary valve regurgitation. Normal tricuspid valve. Mild tricuspid valve regurgitation..." | 1. The report mentions 'Moderate tricuspid valve regurgitation' and 'Moderate aortic valve regurgitation', which indicates the presence of moderate regurgitation in at least one of the valves.<br>2. Since the report does not explicitly mention the degree of mitral regurgitation, but it does mention 'Trivial mitral valve regurgitation', we can infer that the mitral valve regurgitation is not absent or trivial.<br>3. Given the presence of moderate regurgitation in other valves and the absence of 'absent' or 'trivial' for mitral valve regurgitation, the **most fitting answer is 'mild to moderate'.** | Trivial MR | Mild to Moderate MR |
| Not following the prompt instructions | "...Status post 27/29 mm [...] mechanical bileaflet aortic valve prosthesis [...]. Trivial aortic valve prosthetic regurgitation (washing jets). Aortic valve prosthesis systolic mean Doppler gradient 12 mmHg. Aortic valve prosthetic orifice area by Doppler: 2.29 cm2 Thickened mitral valve. Trivial mitral valve regurgitation. Normal pulmonary valve. Trivial pulmonary valve regurgitation. **Normal tricuspid valve. Trivial tricuspid valve regurgitation...**" | 1. The report explicitly mentions 'Normal tricuspid valve.'<br>2. The report does not mention 'trivial,' 'mild,' 'mild to moderate,' 'moderate,' 'moderate to severe,' 'severe,' or 'not specified' tricuspid stenosis.<br>3. Since 'normal' implies no abnormalities, the absence of any mention of tricuspid stenosis leads to the conclusion of 'absent. | Not Specified TS | Absent TS |
| | "...Trileaflet aortic valve. Thickened aortic valve. **Trivial aortic valve regurgitation.** Mitral valve posterior leaflet prolapse. Possible flail mitral valve posterior (P2) leaflet [...] Severe mitral valve regurgitation. Eccentric mitral valve regurgitation directed anteriorly..." | 1. The report mentions 'Trivial aortic valve regurgitation' which indicates a regurgitation degree of 'trivial'.<br>2. Since 'trivial' is not an option and the report does not explicitly mention 'absent' or 'moderate to severe', the only remaining option is 'mild to moderate'. | Trivial AR | Mild to Moderate AR |
| Confabulating nonfactual content | "...Bicuspid aortic valve. Aortic valve systolic mean Doppler gradient 26 mmHg. Aortic valve area by Doppler 2.48 cm2**. Moderate-severe aortic valve regurgitation.** Eccentric aortic valve regurgitant jet. Quantification with PISA was attempted but not reported. Normal mitral valve. Trivial mitral valve regurgitation. ..." | 1. The report mentions 'moderate aortic valve regurgitation'. 2. The term 'moderate' implies a degree between 'mild' and 'severe'. 3. Since 'moderate' is not an option, the closest appropriate category is 'mild to moderate'. Final Answer: mild to moderate | Moderate To Severe AR | Mild to Moderate AR |

| | | | | |
|---|---|---|---|---|
| | "Status post [...] mm aortic valve [...] prosthesis. **Thickened or calcified aortic valve tissue prosthesis with nodular calcification and degenerative strands** ... Normal mitral valve. Trivial mitral valve regurgitation. **Thickened tricuspid valve. mild to moderate tricuspid valve regurgitation...** " | 1. The report mentions a **prosthetic tricuspid valve.**<br>2. The prosthetic valve is described as having thickened tissue with degenerative strands.<br>3. There is no mention of a prosthetic valve, but rather a degenerative condition of the native valve. | No Prosthetic Tricuspid Valve | Prosthetic Tricuspid Valve |

3. **Supplementary Figures and tables:**

| | Valvular Heart Disease | Not specified | Absent | Trivial | Mild | Mild to Moderate | Moderate | Moderate to severe | Severe | Prosthetic Valve |
|---|---|---|---|---|---|---|---|---|---|---|
| **Prompt Optimization Set (n=200)** | | | | | | | | | | |
| Aortic Valve | Regurgitation | 2.5 | 45.5 | 25.0 | 9.0 | 5.0 | 1.0 | 0.5 | 1.5 | 10 |
| | Stenosis | 82.0 | 0.5 | 0 | 3.5 | 1.0 | 0.5 | 1.0 | 1.5 | |
| Mitral Valve | Regurgitation | 2.0 | 5.0 | 52.5 | 24.0 | 9.0 | 2.5 | 1.5 | 0 | 3.5 |
| | Stenosis | 94.0 | 0 | 0.5 | 0.5 | 0.5 | 0.5 | 0.5 | 0 | |
| Pulmonary Valve | Regurgitation | 20.5 | 5.5 | 55.0 | 18.5 | 0 | 0.5 | 0 | 0 | 0 |
| | Stenosis | 99.5 | 0.5 | 0 | 0 | 0 | 0 | 0 | 0 | |
| Tricuspid valve | Regurgitation | 3.5 | 0.5 | 43.5 | 34.0 | 7.0 | 7.5 | 1.5 | 2.5 | 0 |
| | Stenosis | 100 | 0 | 0 | 0 | 0 | 0 | 0 | 0 | |
| **Test Set (n=1000)** | | | | | | | | | | |
| Aortic Valve | Regurgitation | 1.3 | 51.3 | 21.1 | 12.2 | 2.8 | 1.0 | 0.6 | 0 | 9.7 |
| | Stenosis | 82.3 | 0.3 | 0.9 | 2.4 | 0.5 | 1.5 | 0.4 | 2.0 | |
| Mitral Valve | Regurgitation | 1.6 | 3.8 | 56.8 | 21.2 | 9.3 | 3.4 | 1.1 | 1.5 | 1.3 |
| | Stenosis | 97.4 | 0.1 | 0 | 0.2 | 0.1 | 0.1 | 0.5 | 0.3 | |
| Pulmonary Valve | Regurgitation | 22.1 | 3.0 | 63.9 | 9.1 | 1.1 | 0.3 | 0 | 0.1 | 0.4 |
| | Stenosis | 99.3 | 0.3 | 0 | 0 | 0 | 0 | 0 | 0 | |
| Tricuspid valve | Regurgitation | 1.8 | 0.7 | 53.5 | 25.8 | 10 | 3.6 | 1.5 | 2.5 | 0.6 |
| | Stenosis | 99.4 | 0 | 0 | 0 | 0 | 0 | 0 | 0 | |

**Table S1.** Prevalence of prosthetic valves and valvular heart disease severities in the test dataset.

| | Model (instruct) | AR | AS | MR | MS | PR | PS | TR | TS | Prosthetic Aortic (n=97) | Prosthetic Mitral (n=13) | Prosthetic Pulmonary (n=4) | Prosthetic Tricuspid (n=6) | Mean VHD Severity | Mean Prosthetic Valves |
|---|---|---|---|---|---|---|---|---|---|---|---|---|---|---|---|
| **Accuracy** | Qwen2-72B | 99.4 | 95.6 | 99 | 100 | 98.5 | 100 | 98.5 | 98 | 100 | 100 | 100 | 100 | 98.6 | 100.0 |
| | Llama3-70B | 99.4 | 100 | 99 | 97.4 | 98.5 | 100 | 98 | 100 | 100 | 100 | 100 | 100 | 99.0 | 100.0 |
| | Mixtral8 | 84.7 | 83.5 | 92.8 | 60.2 | 87.4 | 98.9 | 81.5 | 97.9 | 98 | 94.5 | 91.5 | 98 | 85.9 | 95.5 |
| | Llama3-8B | 79.4 | 65 | 90.5 | 7.4 | 96.5 | 4 | 87.5 | 81.5 | 100 | 98 | 100 | 100 | 64.0 | 99.5 |
| | Phi3-mini | 51.7 | 50 | 56.8 | 50.5 | 48.7 | 86.4 | 34.7 | 54.1 | 97.5 | 98.5 | 99.5 | 98 | 54.1 | 98.4 |

| | Model (instruct) | AR | AS | MR | MS | PR | PS | TR | TS | Mean VHD Severity |
|---|---|---|---|---|---|---|---|---|---|---|
| **Mean Squared Error** | Qwen2-72B | 0.05 | 0.91 | 0.04 | 0 | 0.04 | 0 | 0.01 | 0.07 | 0.14 |
| | Llama3-70B | 0.05 | 0 | 0.04 | 0.03 | 0.04 | 0 | 0.12 | 0 | 0.04 |
| | Mixtral8 | 0.33 | 1.73 | 0.09 | 1.76 | 0.48 | 0.1 | 0.51 | 0.14 | 0.64 |
| | Llama3-8B | 0.35 | 3.52 | 0.13 | 7.3 | 0.14 | 7.5 | 0.45 | 1.1 | 2.56 |
| | Phi3-mini | 0.97 | 3.22 | 1.65 | 1.22 | 1.98 | 0.36 | 2.95 | 0.46 | 1.60 |

**Table S2.** Mean accuracy of language models in detection of severity degree and prosthetic presence.

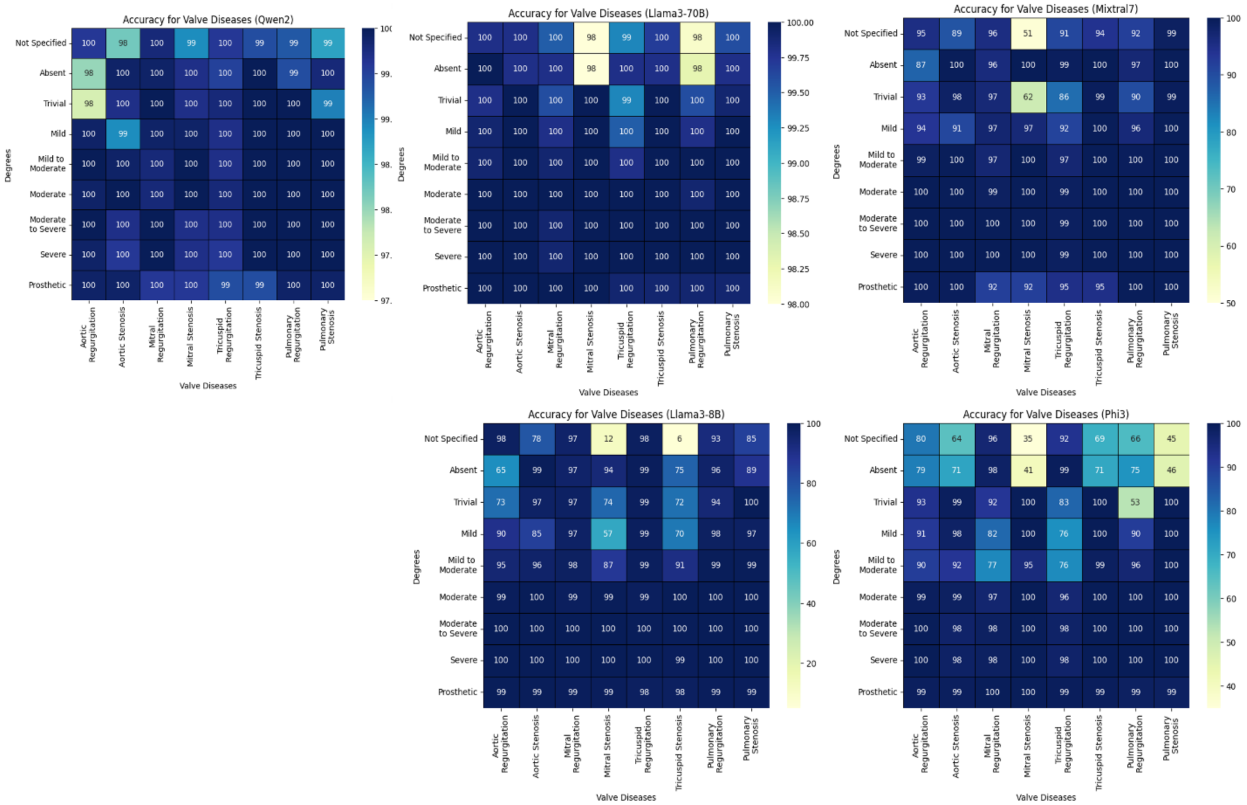

**Figure S2.** Classification matrix presenting accuracy of five models in the detection of 8 valvular diseases and 8severity degrees + prosthetic valve presence

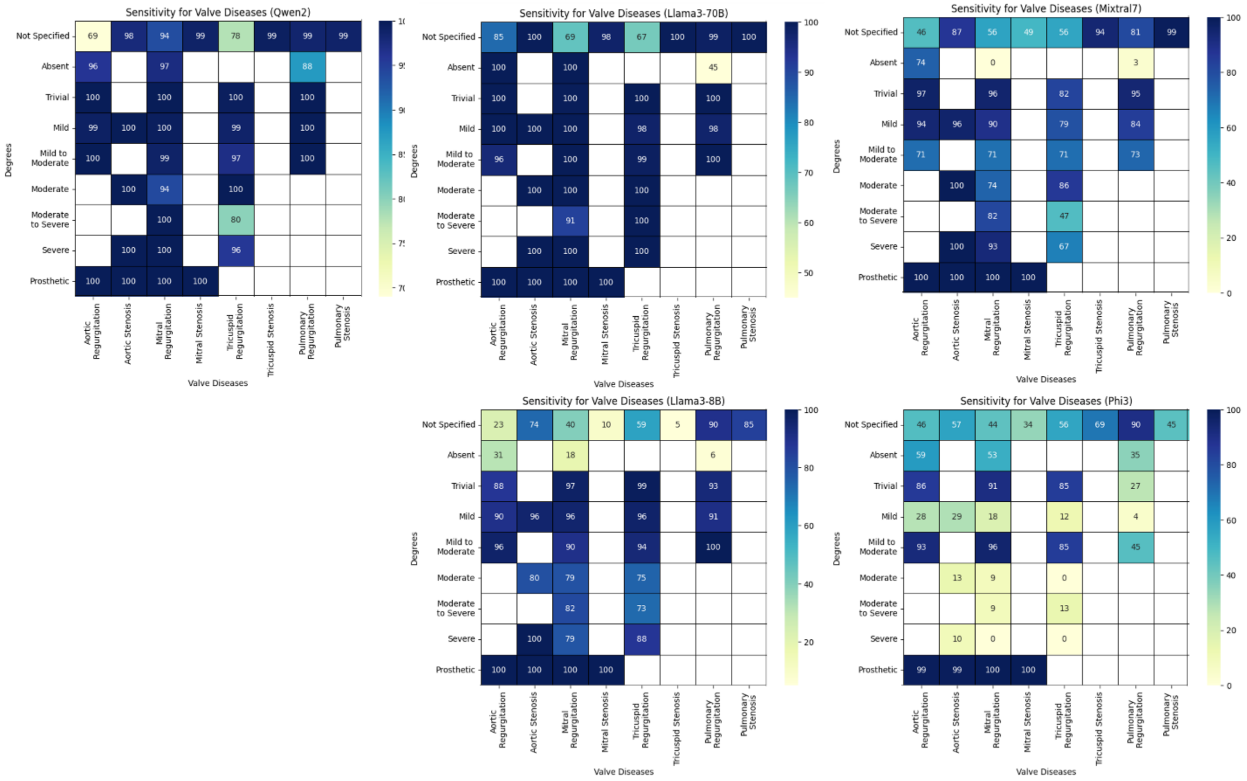

**Figure S3.** Classification matrix presenting sensitivity of five models in the detection of 8 valvular diseases and 8severity degrees + prosthetic valve presence

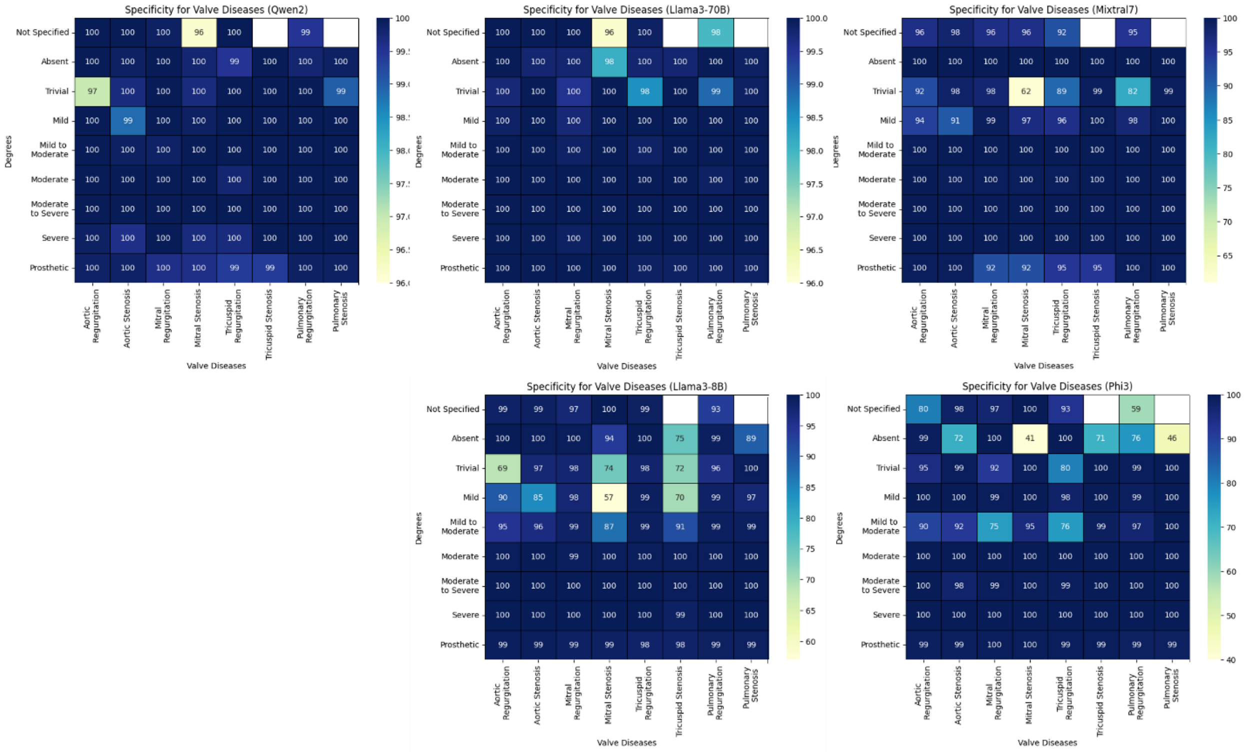

**Figure S4.** Classification matrix presenting Specificity of five models in the detection of 8 valvular diseases and 8severity degrees + prosthetic valve presence

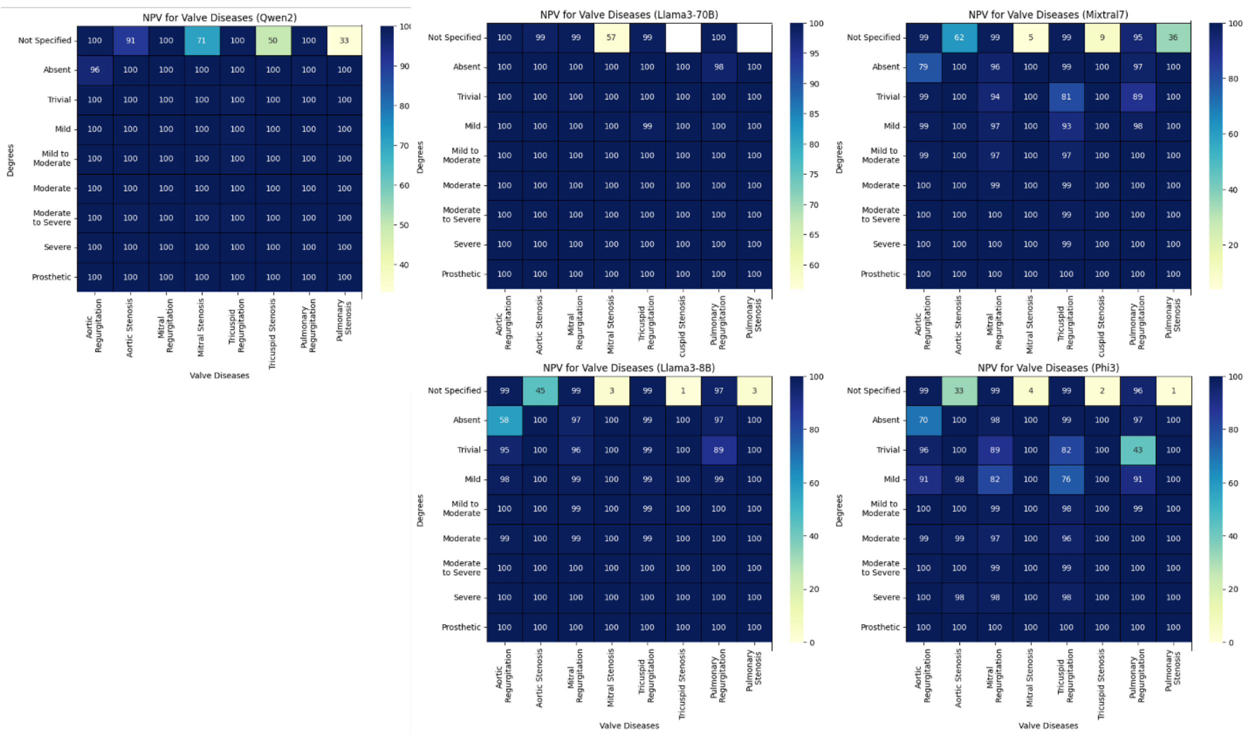

**Figure S5.** Classification matrix presenting negative predictive value (NPV) of five models in the detection of 8 valvular diseases and 8severity degrees + prosthetic valve presence

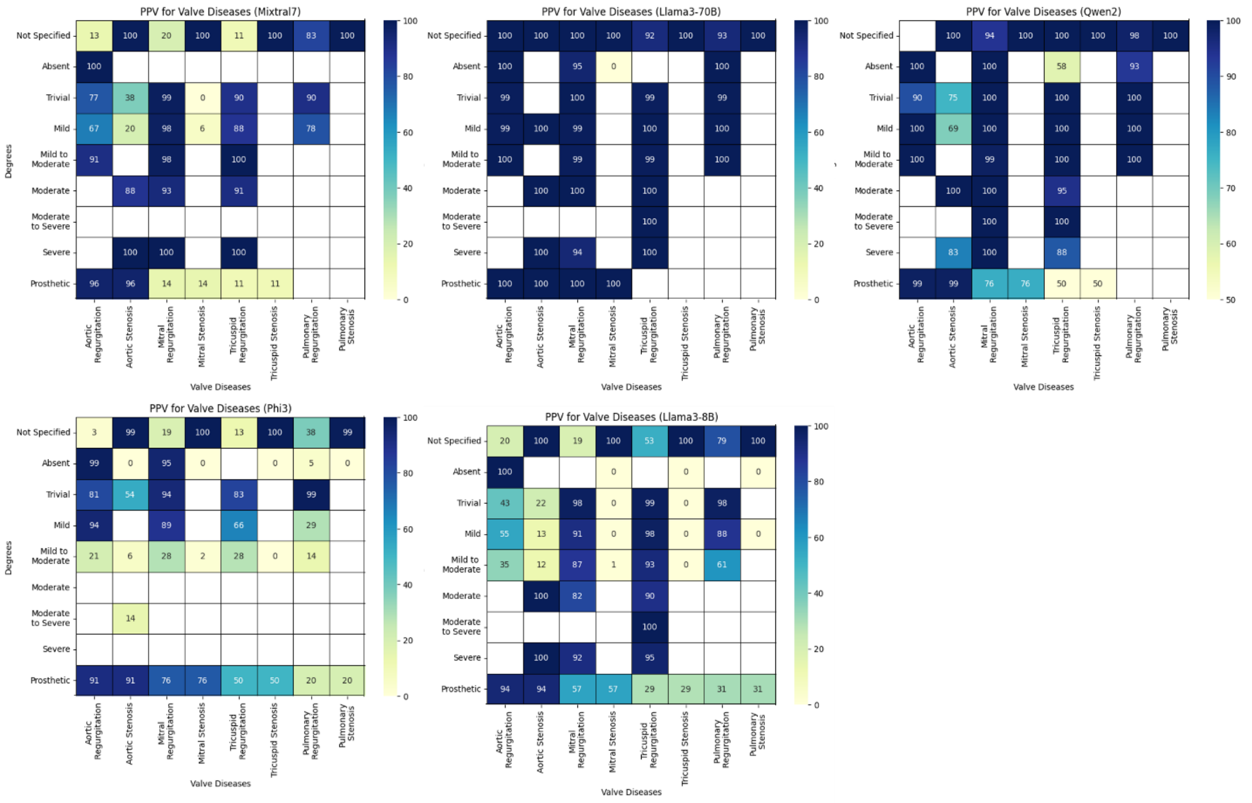

**Figure S6.** Classification matrix presenting positive predictive value (PPV) of five models in the detection of 8 valvular diseases and 8severity degrees + prosthetic valve presence

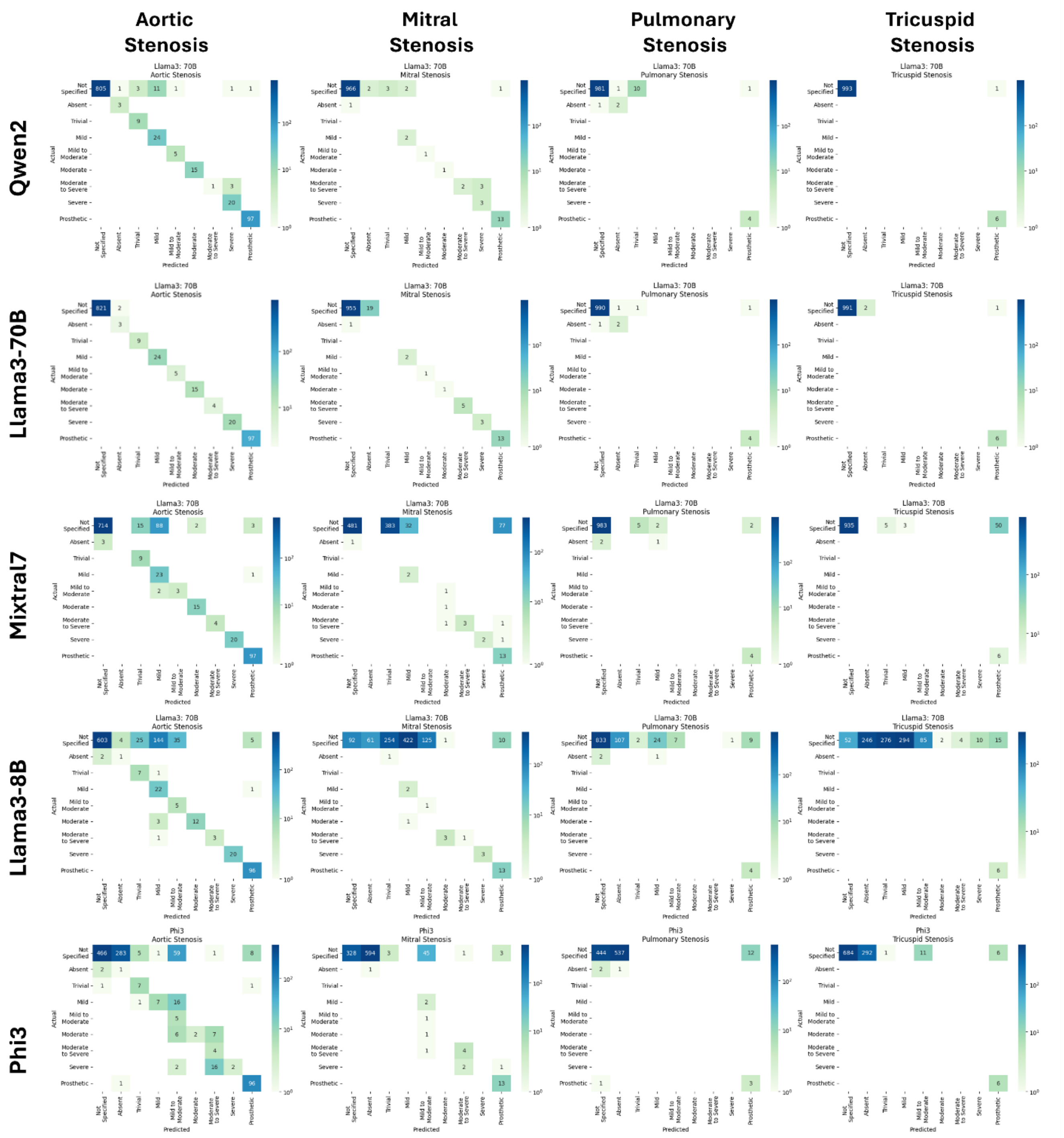

**Figure S7.** Heatmaps presenting five models' predicted labels versus actual labels for four stenotic valvular heart diseases
